# A health economic evaluation of calorie labelling in the out-of-home sector in England: A modelling study

**DOI:** 10.64898/2026.01.19.26344381

**Authors:** Penny Breeze, Katharine Pidd, Alexandra Kalbus, Laura Cornelsen, Kerry Ann Brown, Steven Cummins, Dalya Marks, Cherry Law, Richard Smith, Oana-Adelina Tanasache, Vanessa Er, Camilla Forbes, Alan Brennan

## Abstract

**Objective:** In England, since 2022, large businesses providing food in the out-of-home sector are required to display calorie information for non-prepacked food and non-alcoholic drink items. This study estimates long-term cost-effectiveness of the policy by extrapolating real-world evidence on short-term policy effects in England.

**Design:** The lifetime health economic impacts of calorie labelling were simulated using a microsimulation model. The analysis adopted a health systems perspective to compare the policy with a counterfactual ‘no intervention’ scenario. The policy may impact calories consumed through consumer behaviour changes and through menu changes based on observations from real-world evaluations. Estimated changes to daily calorie intake are translated to weight changes. Simulated outcomes include changes in obesity, diabetes cases, cardiovascular events, quality adjusted life years (QALYs) and National Health Service costs with probabilistic sensitivity analysis to describe uncertainty.

**Setting:** A synthetic population for England aged 13–79 combining data was generated the National Diet and Nutrition Survey (2009–19) and Health Survey for England (2018, 2019).

**Participants:** None

**Results:** The policy is estimated to generate lifetime cost savings were estimated to be-£9.15 (95% CI - £31.63, £2.50), and incremental QALYs 0.0021 (95% CI-0.0008, 0.0048) per person. The incremental net benefit at £20,000 per QALY was £50.23 (95% CI-£16.41, £74.68). Greater cost-savings and QALY gains were observed in the most deprived groups.

**Discussion:** The out-of-home calorie labelling policy in England is uncertain but most likely cost-effective with cost-savings and marginally beneficial to health. The results are driven by expected menu changes.

## Introduction

Obesity has increased globally in recent years and in 2022 to 2023, 64.0% of adults aged 18 years and over in England were estimated to be overweight or living with obesity (1, 2). Obesity is associated with reduced life expectancy and is a risk factor for a range of chronic diseases, including cardiovascular disease and type 2 diabetes mellitus (3). The social and economic consequences of obesity are concerning for governments with a strong association with socioeconomic position (2) and high economic impact (4). There is a compelling argument for concerted action to address the rising prevalence of obesity, improve population well-being and benefit society. One such action is to target food purchased in the out-of-home sector, as this setting is associated with increased dietary intakes of energy and nutrients of concern, e.g. saturated fat, salt and sugar (5).

In April 2022, the government introduced a mandatory calorie labelling policy in England, which required large businesses (with 250 or more employees) selling food and drinks (e.g. restaurants, cafes, fast-food chains) to show calorie information (in kilocalories(kcal)) of food and non-alcoholic drink items prepared for immediate consumption at the point of choice (6). Prior to implementation, studies suggested that only 17–24% of businesses provided in-store calorie labelling (7, 8). The objective of the calorie labelling regulations was to support consumers to make informed choices by providing calorie information, and encourage businesses to reformulate their menus to reduce the calorie content of the food on offer (6).

Evidence for the effect of calorie labelling on calories purchased and consumed in the out-of-home sector reports mixed findings. A meta-analysis of studies found that menu labelling alone was not significantly effective in modifying menu selection (-37kcal, 95% CI-95.85 to 34.18), but was effective if accompanied by information to add context and interpretation (-67kcal 95% Confidence Interval (CI)-116.99 to-17.79) (9). Alternative meta-analysis of calorie labelling in out of home settings estimate statistically significant reductions of-47 and-11 kcal in energy purchased per 600 kcal meal (10, 11). Furthermore, meta-analysis estimated that retailers reduced the energy content of menu items by 15 kcal on average (95% CI 8 to 23) following calorie labelling (12).

Mathematical models can be used in public health to bring together information from multiple sources to estimate policy effects (13). Models provide policymakers with important evidence to assess the effectiveness and cost-effectiveness of policy interventions by extrapolating short-term changes in intermediate outcomes, such as calorie changes, to long-term outcomes such as type 2 diabetes and cardiovascular diseases (14). Furthermore, disease risks and responsiveness to the policy can vary between socioeconomic groups. Heterogeneous effects can be explored with simulation modelling to identify social groups that benefit more from policies.

Calorie labelling policies in the out-of-home sector have been evaluated in modelling studies for the United Kingdom and European countries (15, 16), and cost-effectiveness analyses for the United States (17–19) and Kenya (20). All studies found that the calorie labelling policy would improve long term health outcomes. All studies analysed the impact of changes in consumer behaviour, and four incorporated effects from menu reformulation (15–17, 19).

However, implementing public health policy in a complex and dynamic food system adds uncertainty to the effectiveness of the calorie labelling policy, and economic evaluations should be updated with new real-world evidence that reflects the policy effect within the context at the time of implementation (21). Real-world evaluations of the policy, using natural experimental methods, capture aspects of implementation, such as compliance, enforcement, and consumer awareness (22, 23), that impact the effectiveness of the calorie labelling policy. Evidence from real-world evaluations of the calorie labelling policy in England have not identified a statistically significant impact on calorie consumption (24). However, there is some evidence of reduction in the calorie content of food available through menu changes in the out-of-home sector since the introduction of the policy (25, 26).

The aim of this study is to estimate the lifetime cost-effectiveness of the out-of-home calorie labelling policy in England on health outcomes and healthcare costs, and to observe the distribution of these impacts across socioeconomic groups. The analysis is novel as it draws on emerging evidence from real-world evaluations of the policy to understand the effects of the intervention.

## Method

### Health Economic Analysis

We evaluated the lifetime discounted costs and Quality-Adjusted Life Years (QALYs) to estimate the incremental net benefit of calorie labelling compared with no intervention assuming a willingness-to-pay threshold of £20,000 per QALY (27). A lifetime perspective was chosen to capture long-term impacts of body mass index (BMI) on non-communicable disease outcomes and mortality. The analysis adopted a health system perspective to include direct medical costs, social care costs and costs incurred by actors whose primary intent is to improve human health (28). In this example the analysis includes the cost to national and local governments to implement and enforce the calorie labelling policy, but excludes costs incurred by businesses. Costs were inflated to 2022 prices, and costs and QALYs were discounted at a rate of 3.5% (27). The analysis considered the outcomes combining effects on consumers and menu changes which include reformulation or introducing new dishes. The scenario is compared with a counterfactual scenario in which no intervention is implemented. We conducted probabilistic sensitivity analysis (PSA) with 1,000 samples taken from an appropriate statistical distribution. A Consolidated Health Economic Evaluation Reporting Standards 2022 (CHEERS) checklist for the analysis is provided in the supplementary material section 11 (29).

### Baseline Population

The baseline population was based on individuals aged 13–79 extracted from the Health Survey for England (HSE) 2018 and 2019 (30). From this dataset, information on individual’s’ age, sex, ethnicity, socioeconomic profile (Index of Multiple Deprivation and National Statistics Socio-economic classification (NS-SEC)), metabolic risk factors (weight, height, systolic blood pressure, total cholesterol, HDL cholesterol, HbA1c), medical history and health-related quality of life (EQ-5D) was collected. Missing data were imputed to preserve demographic representation in the dataset, using predictive mean matching. Age in years was imputed using additional data from HSE 2014. The National Diet and Nutrition Survey (NDNS) collects detailed information on food consumption in the UK (31). The survey asks respondents to report on the frequency that respondents purchase meals out-of-home and from take-aways.(31). Data synthesis was conducted to combine individual data from the HSE and NDNS accounting for common sociodemographic factors including age, sex, NS-SEC and ethnicity. The method uses iterative proportional fitting (32) and matches individuals from HSE to out-of-home eating preferences within similar sociodemographic groups reported in the NDNS population. Table 1 summarises the characteristics of the synthetic population with details on data synthesis provided in section 2 of the supplementary material.

**Table 1:** Estimated change in average calories from behavioural change and menu changes in base case.

|  | Mean | 95% Confidence Interval | Source |
| --- | --- | --- | --- |
| Base case change in calories per day for behavioural effects | -2 | -35.9 to 33.1 | Kalbus et al. (33) |
| Base case change in calories per dish on menu | -9 kcal | -1 to -16 | Essman et al (25) |
| Base case times per week purchase occasions per week from out-of-home sector | 1.56 | NA | Author's own analysis * |
| Year 1 implementation cost | £25,000 | NA | Impact Assessment (35) |
| Ongoing enforcement costs (per year) | £100,000 | NA | Impact Assessment (35) |
| SA 3: Male and low SES purchase occasions per week from out-of-home sector | 1.50 | NA | Author's own analysis * |
| SA 3: Male and high SES purchase occasions per week from out-of-home sector | 1.65 | NA | Author's own analysis* |
| SA 3: Female and low SES purchase occasions per week from out-of-home sector | 1.43 | NA | Author's own analysis* |
| SA 3: Female and high SES purchase occasions per week from out-of-home sector | 1.57 | NA | Author's own analysis* |
| SA 4: IMD1 (most deprived) | -20.22 | -21.44 to -18.99 | Unpublished analysis (Kalbus et al. (33) Calorie labelling and changes in the calorie content of food and drink available for online food delivery over time) |
| SA 4: IMD2 | -19.07 | -20.43 to -17.70 |  |
| SA 4: IMD3 | -16.25 | -17.82 to -14.67 |  |
| SA 4: IMD4 | -17.13 | -18.81 to -15.45 |  |
| SA 4: IMD5 (least deprived) | -16.23 | -17.86 to -14.60 |  |
| SA 7: change in calories per dish for behavioural effects | -47. |  | Crocket et al. (10) |
| SA 7: change in calories per dish on menu | -15 |  | Zlatevska et al. (12) |

\*Authors' own analysis Kantar's Worldpanel OOH Purchase panel, 47w/e, 27th Nov 2022

### Intervention effects

The intervention effects and cost of calorie labelling are reported in Table 2. The effect of the calorie labelling policy on calories is described through two mechanisms. First, calorie labelling may impact consumer behaviour, i.e. the choice of foods from menus affected by the policy.

**Table 2:** Simulated baseline population representative of the general population of England aged 13-79 (N=200,000)

|  | % |  |
| --- | --- | --- |
| Male | 49.02 |  |
| Female | 50.98 |  |
| Age 13–24 years | 17.39 |  |
| Age 25–34 years | 16.75 |  |
| Age 35–44 years | 16.37 |  |
| Age 45–54 years | 16.09 |  |
| Age 55–64 years | 15.85 |  |
| Age 65–79 years | 16.49 |  |
| IMD5 (least) | 18.83 |  |
| IMD4 | 19.59 |  |
| IMD3 | 19.83 |  |
| IMD2 | 20.89 |  |
| IMD1 (most) | 20.86 |  |
| Normal weight^ | 38.00 |  |
| BMI 25-30kg/m<sup>2</sup> ^ | 34.58 |  |
| BMI >=30kg/m<sup>2</sup> ^ | 27.42 |  |
| White ethnicity | 84.53 |  |
| Non-white ethnicity | 15.47 |  |
| Out-of-home consumption (takeaways or meals out) | 84.75 |  |
|  | Mean | Standard deviation |
| Age | 44.59 | 18.27 |
| BMI | 27.33 | 5.90 |
| IMD Index of Multiple Deprivation, BMI Body Mass Index |  |  |
| ^ Estimated for sample over age of 18 only |  |  |

Second, restaurants may change their menus (recipe change, portion size or new product) in response to the policy. In the base case, both mechanisms are implemented in the model in combination to account for uncertainty in the effectiveness across each component. We compare the intervention with a counterfactual ‘no intervention’ scenario.

Consumer behaviour change estimate was taken from a post-implementation study using controlled interrupted time series analysis of market research data for the out-of-home sector. This study estimated a non-significant reduction in calories purchased of-31.2 kcal (95% CI - 351.8 to 289.4), and an increase in calories in the time trend coefficient (33). The total change in calories purchased per day was estimated based on the difference in calories observed in the first six months after the policy was implemented. The analysis assumes equivalence in purchases and consumption of calories. The time trend estimate in this analysis indicated that any initial change in calorie consumption was not sustained over the long term, a pattern also seen in other studies (34). We assume consumer behaviour returns to baseline levels after six months, with the total change in calories applied over the first cycle of the model as a daily reduction in calories. We estimated an average daily reduction in calories of 2 kcal per day (95% CI-36.4 to 32.6) which suggests that the policy is less effective than previous analyses of consumer responses to purchases and this parameter input was tested in sensitivity analysis (10), but similar in magnitude to an updated meta-analysis (11). Within PSA samples calorie change could increase, or decrease, from baseline due to uncertainty in the parameters sampled, allowing effects to lead to weight loss or weight gain. Due to the small and non-significant effects on consumer behaviour, we also assume no impact on calories consumed across other meals.

Changes in restaurants’ response to the policy are captured through the estimated change in average calories on the menu pre-vs. post-policy in England. Essman et al. (25) estimated that the average calories per menu item was reduced by 9 kcal (95% CI 16 to 1 kcal). Daily calorie change per person was then estimated by multiplying the change in average calories per menu item by the number of dishes purchased per week (1.56 per week) (authors’ analysis of Kantar’s Worldpanel OOH Purchase panel, 47w/e, 27th Nov 2022). Studies from other policy evaluations outside of England estimated a pooled reduction of 15 kcal on average (95% CI 8 to 23) following labelling across multiple studies, which was tested in sensitivity analysis (12). Change in daily calories consumed due to menu changes is assumed to be maintained over time, which would lead to a long-term reduction in body weight.

The costs of enforcing the policy are taken from a government policy impact assessment which estimated £25,000 for implementation, and £100,000 per year for enforcement (35). In line with a healthcare provider perspective industry costs for reformulation and menu changes are not included. Table 2 reports the treatment effects and costs, with full details on input parameters and uncertainty distributions used in probabilistic sensitivity analysis provided in section 9 of the supplementary material.

### Simulation model

We used a discrete microsimulation model with one-year cycles that meets the requirements of behavioural interventions for obesity to estimate the effects of the mandatory calorie menu labelling policy in England (36, 37). The model code for the analysis is available on request to the corresponding author. The model sampled 200,000 individuals using population weights generated to match the age sex distribution of the target population. Stability assessment indicated that incremental net benefit was stable above 100,000 individuals for the incremental net benefit, but a larger sample size was required to achieve stability in short-term estimates (Figure S2 supplementary material). Changes to daily calorie intake were converted into changes in weight based on an approximation of the Hall et al. models of calorie to weight conversion (38). Natural history weight and height trajectories were simulated using age trends identified within HSE. Heterogeneity in growth rates by age were captured by recording an individual’s position relative to others within their age group at baseline. This point in the weight or height distribution determines their subsequent value based on distribution of other age categories. Long-term trajectories for other metabolic risk factors (HbA1c, systolic blood pressure, total cholesterol, HDL cholesterol) were simulated based on analyses of existing UK cohort studies and a UK prospective diabetes study following diagnosis with type 2 diabetes (36, 39, 40). Major health events related to obesity and diabetes were simulated conditional on sociodemographic risk factors and BMI, systolic blood pressure, HbA1c and cholesterol as derived from the epidemiological studies. Cardiovascular disease, cancer, dementia, end stage kidney failure, increase the risk of mortality and all-cause mortality was conditional on age and sex based on population life tables (41). Full details of the model, including parameter inputs are provided in the supplementary materials.

The microsimulation assigns costs to healthcare utilisation throughout the life course of the simulated population. GP attendance, diabetes diagnosis, medication and monitoring, hypertension treatment, statin use all incur healthcare costs from recommended UK costing sources (27). Major health events such as cardiovascular disease, cancer, osteoarthritis and dementia incur healthcare costs identified from costing studies (42–44). Social care costs were estimated for individuals aged over 65 years, and were conditional on age and BMI (45).

### Uncertainty analyses

Probability distributions were assigned to parameter inputs to the model. Details on the form of distribution are provided sections 3-9 of the supplementary material. Probabilistic sensitivity analysis describes the impact of parameter uncertainty, and simulation outcomes are reported with 95% credible intervals (CRI). Subgroup analyses were stratified by index of Multiple Deprivation (IMD) quintiles due to ongoing interest in the sociodemographic impacts of the policy (39) and in line with interest from project stakeholders and public participants. Two sensitivity analyses (SA) estimated the impact of consumer behaviour and menu reformulation changes separately (SA1 and SA2, respectively). All other sensitivity analyses (SA 3-11) were conducted with a sample of 100,000 due to computation time. Two sensitivity analyses tested the sensitivity of the results to variation in the effectiveness of the intervention by socioeconomic status (SA3 and SA4). In SA3, the policy effects were stratified by sex and occupational social class to incorporate estimations an analysis of Kantar that predicted larger, but non-significant, behavioural response for high occupational social class women and more frequent out-of-home consumption for high occupational social class men. The analysis was conducted as part of a previous study but not previously reported (see supplementary material) (33). In SA4, menu changes were estimated from changes in average calories between June 2022 and June 2023 on two online delivery service platforms stratified by IMD (based on where restaurants deliver to) to allow for heterogeneity in menu changes (26). Other sensitivity analyses varied discount rates to 0% and 5% (SA5 and SA6). A sensitivity analysis utilised alternative sources of evidence for consumer (SA7-47 kcal per meal (10), SA8-11 kcal per meal (11)) with reformulation effects (-15 kcal per meal). In SA9, a three-year delay in menu change effects was simulated reflecting observations that reformulation is more likely driven by new lower calorie menu items rather than changes to existing items, extending the time for consumers to change their habits (46). SA10 includes an indirect effect on blood pressure, cholesterol and HbA1c via changes in weight (47). Finally, we vary the method for converting calories to weight in SA11 (48).

## Results

The simulation predicts a reduction in obesity (-69,330 cases (95% CRI-613,104 to 569,628) for the English population aged 13-79 in the first 12 months of the policy (Table 3) and modest reductions in diabetes (-3,684 95% CRI-17,175 to 4300), myocardial infarction (-136 95% CRI - 1,808 to 1,582) and cardiovascular deaths (-92 95% CRI-1,582 to 1,130) after 10 years. Table 3 also reports the reduction in obesity for SA1 (behaviour change only:-33,789 95% CRI-592,708 to 613,527) and SA2 (menu reformulation only:-35,597 95% CRI-63843,-14,124). The higher uncertainty in the effects of the policy to change consumer behaviour has a notable impact on the credible intervals for the base case and SA1 scenarios.

**Table 3:** Incremental health impact of policy for the English population aged 13–79 years (N=45,198,512)

|  | Difference in number with BMI>30kg/m<sup>2</sup> after 1 year | Change in cases of diabetes after 10 years | Changes in myocardial infarction events after 10 years | Changes in stroke events after 10 years | Change in cardiovascular mortality after 10 years |
| --- | --- | --- | --- | --- | --- |
| Combined behavioural and menu changes | -69,330<br>(-613,104, 569,628) | -3,684<br>(-17,175, 4,300) | -136<br>(-1,808, 1,582) | -113<br>(-1,582, 904) | -92<br>(-1,582, 1,130) |
| SA1. Behavioural calorie effect | -33,789<br>(-592,708, 613,527) | -360<br>(-9,503, 9,497) | -18<br>(-904, 904) | -18<br>(-621,684) | 2<br>(-910, 904) |
| SA2. Menu changes | -35,597<br>(-63,843, -14,124) | -3,294<br>(-10,853, 452) | -136<br>(-1,582, 1,356) | -100<br>(-1,356, 904) | -91<br>(-1,356, 1,130) |
| 95% Credible Intervals reported within brackets; SA Sensitivity Analysis |  |  |  |  |  |

**Table 3:** The lifetime discounted costs and QALYs for calorie labelling with 95% credible intervals.

|  | Discounted policy cost | Lifetime health care discounted costs | Lifetime discounted QALY | Incremental total costs vs. No intervention | Incremental QALYs vs. No intervention | Incremental Net Monetary Benefit | % chance cost-effective at £20,000 per QALY |
| --- | --- | --- | --- | --- | --- | --- | --- |
| No intervention |  |  |  |  |  |  |  |
| Average per person effects | £0 | £24,220<br>(£16,885, £38,310) | 15.3369<br>(14.9178, 15.6777) | - | - | - |  |
| Combined consumer and industry effects |  |  |  |  |  |  |  |
| Average per person effects | £0.06 | £24,211<br>(£16,847, £38,321) | 15.3290<br>(14.9194, 15.6789) | £-9.15<br>(£-31.63, £2.90) | 0.0021<br>(0.0008, 0.0048) | £50.23<br>(£-16.41, £74.68) | 93.6% |
| SA1: Consumer purchases effects |  |  |  |  |  |  |  |
| Average per person effects | £0.06 | £24,220<br>(£16,853, £38,309) | 15.3327<br>(14.9240, 15.6849) | £-0.19<br>(£-10.51, £10.67) | -0.0002<br>(-0.0025, 0.0008) | £-3.88<br>(£-42.18, £10.46) | 53.9% |
| SA2: Industry reformulation effects |  |  |  |  |  |  |  |
| Average per person effects | £0.06 | £24,211<br>(£16,847, £38,308) | 15.3352<br>(14.9248, 15.6855) | £-8.91<br>(£-27.90, £0.08) | 0.0023<br>(0.0002, 0.0044) | £54.55<br>(£3.48, £73.50) | 98.5% |
| () 95% Credible Intervals SA Sensitivity Analysis QALY Quality-Adjusted Life Years |  |  |  |  |  |  |  |

The base case simulation of mean gain in lifetime QALYs and reduction in lifetime costs results in an incremental net benefit of £50.23 at a willingness-to-pay threshold of £20,000 per QALY (Table 4). Parameter uncertainty in the simulated cost-effectiveness indicates a high probability that the policy is cost-effective in the base case. The scatter plot in Figure 1A highlights that there is uncertainty in the incremental cost-effectiveness estimates, but the majority of the points lie below the £20,000 per QALY threshold.

**Figure 1:**
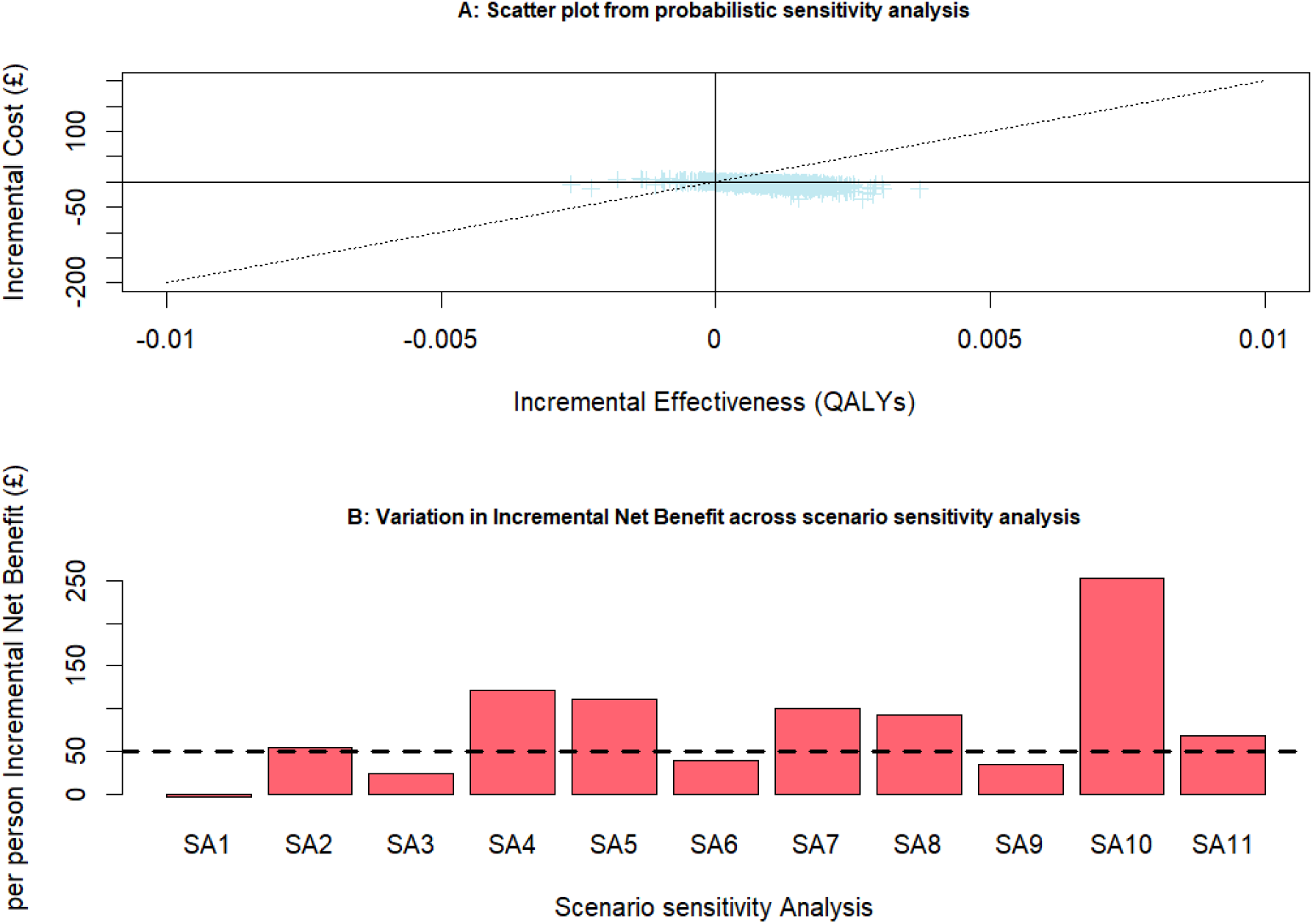
Results of sensitivity analyses (A) Scatter plot showing the results of a probabilistic sensitivity analysis, illustrating the incremental cost versus incremental effectiveness (QALYs). The dashed line represents the willingness-to-pay threshold. (B) Bar chart depicting the variation in per-person incremental net benefit across different sensitivity analyses (SA1 to SA10). The dashed horizontal line indicates the average per-person incremental net benefit for the base case analysis. SA1: behavioural effect only, SA2: menu reformulation effect only, SA3: Effects stratified by Sex and SES, SA4: Reformulation effect by IMD quintile, SA5: Discount rate 0%, SA6: Discount rate 5%, SA7: Meta-analysis intervention effects, SA8: secondary meta-analysis intervention effects, SA9: Delayed reformulation effect, SA10: Metabolic consequence for weight loss, SA11: Alternative calorie to weight equation.

**Table 4:** The lifetime discounted costs and QALYs for calorie labelling with effects stratified by socioeconomic groups for the base case and two sensitivity analyses varying the intervention effect.

|  | Discou<br>nted<br>policy<br>cost | Lifetime<br>health care<br>discounted<br>costs | Lifetime<br>discounted<br>QALY | Incremental<br>total costs | Incremental<br>QALYs | Incremental<br>net benefit |
| --- | --- | --- | --- | --- | --- | --- |
| No intervention |  |  |  |  |  |  |
| IMD5 (least deprived) | 0 | 25453 | 15.2561 |  |  |  |
| IMD4 | 0 | 23146 | 15.1280 |  |  |  |
| IMD3 | 0 | 24243 | 15.0859 |  |  |  |
| IMD2 | 0 | 24610 | 15.6450 |  |  |  |
| IMD1 (most deprived) | 0 | 26487 | 15.5074 |  |  |  |
| Combined consumer behaviour and menu reformulation effects |  |  |  |  |  |  |
| IMD5 (least deprived) | 0.06 | 25446 | 15.2579 | -7.54 | 0.0019 | 44.91 |
| IMD4 | 0.06 | 23137 | 15.1300 | -8.35 | 0.0020 | 48.01 |
| IMD3 | 0.06 | 24234 | 15.0980 | -9.34 | 0.0021 | 51.98 |
| IMD2 | 0.06 | 24600 | 15.6472 | -9.65 | 0.0021 | 52.34 |
| IMD1 (most deprived) | 0.06 | 26476 | 15.5096 | -10.62 | 0.0022 | 53.76 |
| SA3. Intervention effect adjusted for SES and sex |  |  |  |  |  |  |
| IMD5 (least deprived) | 0.06 | 25444 | 15.2565 | -8.92 | 0.0004 | 16.92 |
| IMD4 | 0.06 | 23136 | 15.1286 | -9.20 | 0.0006 | 20.79 |
| IMD3 | 0.06 | 24233 | 15.0967 | -9.81 | 0.0008 | 24.94 |
| IMD2 | 0.06 | 24600 | 15.6458 | -9.84 | 0.0008 | 26.06 |
| IMD1 (most deprived) | 0.06 | 26476 | 15.5084 | -10.58 | 0.0010 | 31.10 |
| SA4. Combined consumer behaviour and menu reformulation effects, with alternative reformulation stratified by IMD |  |  |  |  |  |  |
| IMD5 (least deprived) | 0.06 | 25388 | 15.2538 | -14.92 | 0.0039 | 87.68 |
| IMD4 | 0.06 | 23475 | 15.1408 | -18.45 | 0.0046 | 110.35 |
| IMD3 | 0.06 | 24459 | 15.0926 | -19.45 | 0.0047 | 112.55 |
| IMD2 | 0.06 | 24571 | 15.6078 | -24.11 | 0.0055 | 109.24 |
| IMD1 (most deprived) | 0.06 | 26704 | 15.4714 | -28.86 | 0.0061 | 150.90 |
| () Credible Intervals SA Sensitivity Analysis QALY Quality Adjusted Life Years IMD Index of Multiple Deprivation |  |  |  |  |  |  |

In SA1, the behavioural effects of the intervention are found to reduce expected lifetime healthcare costs by £0.19 per person, and reduce expected lifetime QALYs with considerable uncertainty (Table 4). The overall effect on the incremental net benefit is negative, suggesting that the policy is not cost-effective based on expected behavioural impacts alone. Negative QALYs were attributed to non-linearity observed in the model, in which weight gain generates greater losses to QALYs than an equivalent weight loss. If menu change effects are simulated in isolation (SA2), the analysis estimates £9 reduction in lifetime costs per person, and 0.0021 gains to incremental lifetime QALYs.

Stratification of the policy effects by socioeconomic status (Index of Multiple Deprivation) indicates that the lifetime effects on health and heath care cost savings are greater in the most deprived groups (Table 4). As such, in the base case, the policy is not predicted to increase health inequalities after accounting for the greater out-of-home consumption among less deprived and greater risk of disease in less deprived individuals. In SA3 we observe that stratifying effects by socioeconomic position did not increase inequalities in the lifetime health economic outcomes, which is mainly due to the dominating effects of menu changes on lifetime benefits. In SA4 an alternative source of evidence was used for menu reformulation effects, stratified by IMD quintiles. In this analysis, the overall benefits are larger than the base case, with greater effects seen for the most deprived groups. Sub-group analysis by other key characteristics did not identify substantial variation in health economic outcomes (Supplementary material Table S33).

Additional sensitivity results varying individual data inputs and modelling assumptions are reported in supplementary Table S34 and illustrated in Figure 1B. These indicate that the sensitivity analyses report positive incremental net benefit results, with the exception of SA1 which describes the cost-effectiveness if only behavioural effects are incorporated.

## Discussion

This health economic evaluation finds that England’s calorie labelling policy is likely to be cost-effective. This conclusion relies on out-of-home outlets undertaking menu changes resulting in fewer calories on average on offer. If the policy is assumed to only change consumer behaviour in the estimated range, its impact would be too small and uncertain to produce long-term health and economic advantages to offset the costs. On the other hand, even very small changes to calorie intake due to menu changes will generate sufficient health benefits to offset costs.

The long-term benefits from reformulation are expected to be spread across IMD quintiles, with the most deprived IMD groups benefiting most due to greater risks for adverse health events.

This pattern is robust to sensitivity analyses including when effects on behavioural responses differ by high occupational social class, and the effects of reformulation are delayed.

The impact on the prevalence of obesity and mortality are lower than those predicted in a previous modelling study in England (15). Using policy effects from a meta-analysis (10), the authors estimated a-0.31 (95% CR-0·10 to-0·35) percentage point change in obesity over 10 years and 230 (140 to 380) fewer cardiovascular deaths after 10 years (15). We find in our base case an approximately-0.55 (95% CR-4.6 to-4.6) percentage point change in obesity at 12 months and 92 fewer cardiovascular deaths after 10 years. Therefore, there is consistency in the potential health benefit but the uncertainty in the long-term effectiveness of the calorie can have a notable impact on the model results. Our results are consistent with previous cost-effectiveness analyses that report lifetime cost-savings in response to calorie labelling policies, albeit with slightly lower estimates for cost savings (-£9 vs-$68) (25). Previous modelling studies have not reported adverse consequences for health inequalities, when exploring the distribution of benefits across social groups. In our sensitivity analyses we incorporate demographic patterns in consumer behaviour and menu change effects. Despite more affluent groups having larger average responses to the policy, the unequal distribution of underlying health risks for type 2 diabetes and cardiovascular disease in the model compensate for this effect. Therefore, we find greater lifetime QALYs and cost-savings are observed in more deprived IMD quintiles.

Menu change effects are key to the cost-effectiveness of this policy, and yet the evidence supporting our reformulation scenario has limitations. While two studies have detected changes in menu offerings in response to the calorie labelling policy in England, neither can conclude that changes to menus will translate to consumption of less calories consumed. The only real-world study to measure calorie intake before and after policy implementation, accounting for behavioural and reformulation effects, did not report a reduction in calories consumed. However, the study was powered to detect a 7% reduction in calories consumed, which highlights the challenges in detecting small changes in calorie consumption. Two studies have highlighted that menu changes are characterised by the introduction of new lower calorie menu items, rather than a reduction in calorie content of existing products (25, 46). Whereas the post-implementation calorie data collection was conducted 5–9 months after policy implementation, it is possible that reformulation effects may take longer to develop as menu offerings and consumer tastes develop over time. Given the sensitivity of the results to reformulation effects, ongoing evaluation and monitoring of the policy should focus on whether shifts to lower calorie offerings are maintained, and whether consumers adapt tastes to select these options in the long term.

The health economic model translates change in calories to obesity and its complications, but does not consider the broader aspects of health, such as mental health, potential harms for people with disordered eating, and lost productivity (49). Furthermore, the analysis took a healthcare provider perspective, so does not account for the additional costs incurred by industry to prepare for policy implementation. Taking account of these wider societal effects may mitigate some of the benefits of the policy. However, the analysis also does not include the positive societal benefits from avoidance of obesity-related complications, so the overall societal impact is uncertain. We also acknowledge that there are methodological challenges in the design of real-world public health policy evaluation using controlled interrupted time series methods due to underlying limitations in data and study design (33).

The non-significant impact of calorie labelling in the out-of-home sector is consistent with other real-world evaluations in England that did not identify a statistically significant reduction in calorie consumption (22). Despite this, our analysis indicates that even small effects on calorie consumption through menu changes would justify the cost of the policy from a health care perspective. Both our behavioural response and reformulation response are predicted to lead to only a 2kcal reduction in daily calories. Whereas the reformulation effects produce more favourable estimates in the health economic modelling because the changes are maintained over time and the estimate was statistically significant. As a consequence, there is much less uncertainty in the effects. However, the return on investment indicated in this analysis is lower than predictions that used evidence from meta-analyses of studies conducted prior to implementation in England, and suggest that the policy may not be achieving expected health benefits. There are a number of reasons why the policy is having limited effect on calories. First, the policy only requires calorie labelling in large chain outlets, some of which would have introduced calorie labelling voluntarily prior to implementation. Second, evaluation of the policy has shown that post-implementation compliance was found in 80% of eligible outlets in 2022 and only 15% met government guidelines on labelling (23). Previous research has also found that the effectiveness of a calorie labelling policy is sensitive to the prominence and presentation of the calorie information (9). Qualitative evaluation highlights potential explanations for low compliance, including limited customer interest and limited enforcement (22).

Our economic evaluation finds that calorie labelling in the out-of-home sector is likely to be a cost-effective policy, conditional on an assumption that the policy leads to modest menu change in the absence of changes in consumer behaviour. Despite previous concerns that the policy may exacerbate health inequalities, our analysis suggests that the policy is unlikely to worsen health inequalities, and may reduce them if menu changes has a greater effect on lifetime health in deprived areas.

## Data Availability

All data produced in the present work are contained in the manuscript

