## Supplementary Material for "A health economic evaluation of calorie labelling in the out-of-home sector in England: A modelling study"

Supplementary Material – Calorie labelling Policy evaluation

### 1. Model Summary

The aim of the model is to extrapolate the impact of individual dietary changes on health outcomes for the English population 13-79s. Figure 1 shows the logic model outlining the model’s processes and structures. A health economics analysis plan was developed for the study and is available on request from the corresponding author. The analysis plan was shared with the project team and stakeholders prior to the analysis and updated following comments.

*Figure S1: Logic model used to illustrate model pathways and outcomes*

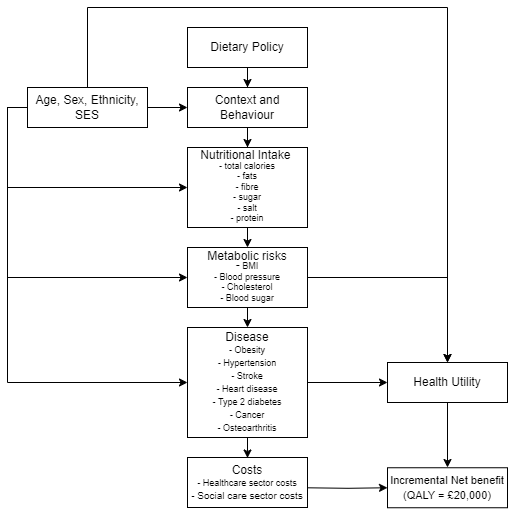

The model uses data from the Health Survey for England (2018,2019) and the National Diet and Nutrition Survey (wave 1-11) to describe individual baseline characteristics and out of home sector consumption patterns. A full description of the process of generating a baseline population is provided in section 2.

### 2. Baseline Population

#### 2.1. Health Survey for England

The model required dietary, demographic, anthropometric and metabolic characteristics that would be representative of the UK general population. The Health Survey for England (HSE) and National Diet and Nutrition (NDNS) were chosen because they collect detailed cross-sectional data on the characteristics of all ages of the English and UK populations. It also benefits from being a reasonably good representation across sociodemographic profiles. A major advantage of the HSE dataset is that it includes important clinical risk factors such as HbA1c, SBP, and cholesterol and health related quality of life (HSE 2018 only). The characteristics of individuals included in the cost-effectiveness model were sampled from the HSE 2018 and HSE 2019 datasets (1, 2). The data were obtained from the UK Data Service.

*Exclusion Criteria*

The total sample size of the HSE 2018 and HSE 2019 were 10,250 and 10,299 respectively. Individuals who were younger than 13 years and older than 79 were excluded from the sample (N=4,915). This left a final sample size of 15,634 individuals.

#### 2.2. National Diet and Nutrition Survey

Data from 11 survey waves of the NDNS comprising a total sample N=15,565, were used in the analysis. The dataset provides a detailed description of individuals’ food and drink consumption, and self-reported consumption habits. The dataset provides a reasonable representation of dietary patterns for the UK due to the survey sampling methods. Given the large variation in individual consumption patterns there are advantages to having a large dataset to capture this heterogeneity, particularly from smaller social groups. The consumption patterns for population sub-samples that may differ from other population groups are important to capture, particularly in order to capture intersectionality in health inequalities for example, minority ethnic groups with young children, or adolescents from deprived socioeconomic backgrounds. Therefore, there are advantages in using all waves from the NDNS data. However, the data have been collected over several years, and earlier surveys may not reflect contemporary tastes and preferences for food. For example, recent shifts away from the consumption of high-sugar soft drinks are not reflected in earlier surveys. A decision was made to use all waves of data in the model to make use of all NDNS waves. It is therefore important to ensure that the analyses are not biased by historical trends in consumption patterns when constructing policy scenarios.

Only individuals between 13 and 79 years of age were included in the sample used to combine information from the HSE and NDNS. We excluded individuals from the sample if they reported a very high or very low calorie intake. Dietary assessments with implausible energy intakes (men: < 800 or > 4200 kcal/days); women: or < 600 or > 3500 kcal/days) were removed from the sample (3). After applying age and calorie intake exclusions, the NDNS sample consisted of N= 9,509. The NDNS sample also provides information on whether individuals consume food in the out-of-home sector.

#### 2.3. Combining HSE with NDNS

Suitable out-of-home data was assigned to each individual within the simulated population based on their socio-demographic characteristics. The two populations, the sampled population using Health Survey for England, and the sampled combined NDNS population, were categorized into 96 mutually exclusive groups based on age, sex and national statistics socio-economic classification 5-level (NSSEC5) (Table S1). These groupings were chosen to balance important determinants of consumption behaviour, while facing limitations in data availability for certain populations. Although ethnicity is a relevant factor, preliminary analyses showed that its inclusion would result in large numbers of sociodemographic groups with very small samples. Ethnicity could only be included at this stage if another variable was removed. To account for the influence of potential determinants of consumption not included in the population groupings, including ethnicity, a two-step procedure was adopted when allocating the NDNS consumption data to the model’s simulated population generated from HSE data. In step 1 individuals are grouped according to the groups identified in Table S1. In step 2 predictive mean matching using imputation methods estimated individuals consumption for one representative variable, using a broader range of demographic variables.

*Table S1: Variable categories used to determine 96 demographic groupings.*

| **Age (years)** | **Sex** | **NS-SEC 5** |
| --- | --- | --- |
| 13 to 15 | Female | Managerial and professional occupations (higher or lower) |
| 16 to 19 | Male | Intermediate occupations |
| 20 to 39 |  | Small employers and own account workers |
| 40 to 69 |  | Low Supervisory and technical occupations |
| 70 or above |  | Semi-routine or routine occupations |
| 13 to 15 |  | Other |

Daily sodium intake was imputed into the modelled population using multiple imputation (predictive mean matching using mice package in R) based on demographic and health data. These included: sex, age category, BMI category, ethnic group, socioeconomic status (NSSEC5), use of Statin or antihypertensive medication, presence of diabetes, fruit and vegetable consumption, housing tenure, and marital status. Following imputation, a nearest-neighbour matching procedure was applied: each individual in the modelled population was matched to an NDNS respondent with a similar sodium intake within the same socio-demographic group. The full dietary profile of the matched NDNS respondent was then assigned to the modelled individual. This approach preserved the correlation structure between different types of food consumption and ensured internal consistency in nutritional data. The initially imputed sodium value was replaced with the matched individual's observed sodium intake, alongside their complete consumption and nutrient profile. Once data on the consumption of sodium was imputed, a nearest neighbour matching process allocated other consumption data for the individual in the NDNS dataset that had the nearest consumption of sodium that were within the same demographic group. The entire consumption data, and out-of-home frequency, of that individual was allocated to ensure that the correlation between consumption types were held, and to ensure consistency in nutritional data. The imputed sodium data was replaced with the selected individual’s consumption of sodium along with full details of consumption and nutritional intake.

#### 2.4. Imputing Missing Data

After the model population has been sampled any missing data is imputed using the mice multiple imputation package in R for missing data relating to weight, hypertensive treatment, diabetes status, systolic blood pressure, total cholesterol, HDL cholesterol and HbA1c. The method is based on Fully Conditional Specification, where each incomplete variable is imputed by a separate model using predictive mean matching.

### 3. Modifying Nutritional Intake and health changes

#### 3.1. Overview of methods

The model has been developed to allow the flexible specification of dietary changes according to the intervention under evaluation. The intervention effects can be applied to all sampled individuals or only a subsample of the population who are assumed to respond to the intervention. Responders can be selected based on demographic characteristics found in the NDNS, for example an age range targeted by the intervention. Similarly, responders may be limited to those who report consuming the targeted product, for example selecting those who report eating in the out of home sector. The model can also apply different intervention effects according to population characteristics.

Once the responding population has been defined the intervention effect can be applied to nutrient intake.

#### 3.2. BMI Change calculator

We have identified 4 economic evaluations of dietary public health policies and interventions in which the Hall et al. equations have been used to describe the relationship between calories and weight change (4-7). Whilst some evaluations have programmed the dynamic weight change equations (5), others have used the equations to approximate the model This method has been adopted in an Australian model for traffic light labelling and junk food taxes In addition, the Department of Health and Social Care calorie model uses this research in their model and assumes that 0.042 kg is lost per calorie reduction A limitation of this simplifying approach is that larger weight losses are expected for a higher initial adiposity, meaning the weight loss in the obese and morbidly obese groups within this model will be underestimated. Furthermore, other factors affecting the relationship, such as age, and sex would be overlooked with this approach.

In the model, weight change is conditional on changes in calories, and the relationship is based on the Hall et al. model. Directly programming the equations into the model would increase the computation burden of the model. A R package”bw” was published, but has been removed from the CRAN repository. For the method to be compatible with the structures, inputs, and computational demands of the SPHR Diabetes Prevention model we approximate the Hall et al. equations. We used the NIDDIK online tool to estimate the impact that other characteristics have on weight changes for a given change in calories. We estimated change in weight conditional on change in calories, % carbohydrate, sodium, and accounting for other characteristics impacting this relationship using the tool. Based on the online tool we estimated the impact that an increase in calories had on weight, and the relationship was slightly stronger for females, higher baseline weight and age. Changes in % carbohydrate and sodium also increase weight. Changes to physical activity and height impact weight change, but these characteristics have not been incorporated into the statistical analysis. We used regression analyses based on the data from the NIDDIK online tool to approximate the impact of changes in calories, sex, age and baseline weight on change in weight. The final parameters and precision of our estimates are reported in Table S2. Interaction terms suggest that the impact of changes in calories increased with age and baseline weight and was less responsive for males.

*Table S2: Estimated change in weight for a given change in calories*

|  | Mean | Standard error |
| --- | --- | --- |
| Change in calories | 0.024161 | 0.0001778 |
| Change in calories by baseline weight | 0.000012 | 1.70e-06 |
| Change in calories by age | 0.000021 | 2.06e-06 |
| Change in calories by male | -0.000941 | 0.000082 |

4. GP Attendance in the General Population

GP attendance was simulated in the dataset to estimate healthcare utilisation for the general population and the costs associated with the number of GP appointments per year, in order to complete a comprehensive cost-effective analysis in the model. A statistical model of GP attendance, conditional on characteristics that are associated, such as age, ethnicity and comorbidities, was developed. If those with life limiting health concerns visit the GP more, then a change in the presence of these conditions will change the number attending GP appointments and thus create an overall change in primary-care costs.

A negative binomial model was used to generate count data and a skewed distribution was observed in the dataset.

$$\mu_{i}=exp(x_{i}\beta)$$

The dispersion parameter of the Negative Binomial distribution $v_{i}$ was sampled from a gamma distribution with mean 1 and variance $\alpha$ based on estimates reported in Table S4. The dose was estimated from the Poisson function.

$$p\left( y>0,x \right)=\frac{\left( v_{i}\mu_{i} \right)^{y}e^{-\left( v_{i}\mu_{i} \right)}}{y!}$$

We used data from the Health Survey for England 2019, which collected a patient self-reported variable on the number times they attended a GP in the last year. It also collected demographic and health related information, including the presence of diabetes. This would allow the GP attendance model to represent the relevant population within the Diabetes Treatment Model. While explicit variables indicating comorbidities were not included, medication data served as proxies. Indicators for prescriptions for antidepressants, lipid lowering medication or anti-hypertensive medication is thought to have a relation with GP attendance, especially considering the policies relating to regular medication reviews. A variable indicating whether an individual has a prescription for these medications is included in the model. Finally, to account for any comorbidities, a variable indicating whether they have a life limiting illness was included. The population included were adults above 20 years old. In the simulation GP attendance for children is not simulated. The characteristics of the study population are reported in Table S3.

*Table S3: Characteristics of HSE 2019*

| Variable | Observations | Mean | SD |
| --- | --- | --- | --- |
| GP Visits | 7,746 | 2.96 | 2.99 |
| Age (20+) | 7,855 | 52.52 | 17.69 |
| Male | 7,855 | 0.46 | 0.50 |
| Black | 7,826 | 0.03 | 0.18 |
| Asian | 7,826 | 0.10 | 0.30 |
| Ethnic Minority | 7,826 | 0.17 | 0.37 |
| BMI | 6,470 | 26.24 | 6.60 |
| Diabetes | 7,848 | 0.08 | 0.27 |
| Life Limiting Illness | 7,847 | 0.24 | 0.43 |
| Lipid Lowering Medication | 4,774 | 0.15 | 0.36 |
| Anti-Hypertensive Medication | 4,768 | 0.24 | 0.43 |
| Antidepressant | 4,774 | 0.10 | 0.30 |

The coefficients of the Negative Binomial model, described in Table S4, were used to calculate the first parameter of the Negative Binomial distribution$\mu_{i}$ based on an individual’s characteristics. The dispersion parameter sampled from the gamma distribution used the alpha reported in Table 6. Three regression specifications are reported in Table S4. Model 1 regresses GP visits against age, sex, ethnicity, BMI, whether they have diabetes and whether they have a life limiting illness. Model 2 regresses GP visits against all parameters in model 1, but with the additional variable indicating if they are prescribed antidepressant medications. Finally Model 3 included whether they are prescribed CVD medication (lipid lowering and anti-hypertensive medication) on top of Model 2.

*Table S4: GP attendance reported in the HSE 2019*

|  | Model 1 | | Model 2 | | Model 3 | |
| --- | --- | --- | --- | --- | --- | --- |
|  | Mean | SE | Mean | SE | Mean | SE |
| Age | 0.003*** | (0.0007) | 0.002** | (0.0009) | -0.002** | (0.001) |
| Male | -0.211*** | (0.0246) | -0.177*** | (0.0295) | -0.220*** | (0.0297) |
| Black | 0.135* | (0.0726) | 0.296*** | (0.0906) | 0.303*** | (0.0902) |
| Asian | 0.211*** | (0.0438) | 0.247*** | (0.0562) | 0.242*** | (0.0557) |
| BMI | 0.011*** | (0.0021) | 0.009*** | (0.0025) | 0.005** | (0.0025) |
| Diabetes | 0.282*** | (0.0438) | 0.238*** | (0.0517) | 0.112** | (0.0533) |
| Life Limiting Illness | 0.700*** | (0.0263) | 0.587*** | (0.0320) | 0.553*** | (0.0319) |
| Antidepressants |  |  | 0.406*** | (0.0417) | 0.407*** | (0.0413) |
| Lipid lowering Medication |  |  |  |  | 0.207*** | (0.0424) |
| Anti-Hypertensive Medication |  |  |  |  | 0.224*** | (0.0392) |
| Constant | 0.422*** | (0.0704) | 0.480*** | (0.0844) | 0.727*** | (0.0883) |
| Alpha | 0.547 | (0.0180) | 0.494 | (0.0206) | 0.476 | (0.0202) |
| LnAlpha | -0.603*** | (0.0330) | -0.705*** | (0.0417) | -0.741*** | (0.0424) |
| N | 6,378 | | 4,215 | | 4,209 | |
| Pseudo R2 | 0.0357 | | 0.0392 | | 0.0434 | |
| Standard errors in parentheses (*** p<0.01, ** p<0.05, * p<0.1) | | | | | | |

The average number of GP visits was approximately 3 times in the year. The number of GP visits was statistically significantly related with all variables included. Individuals that are Black or Asian are more likely to attend the GP, while males are less likely. Those with a higher BMI, a life-limiting illnesses or are taking CVD medication and antidepressants go to the GP more often. The large positive relationships between GP attendance and indicators of ill health justify their inclusion. An unexpected negative relationship is seen in age when including CVD medication variables. This could be explained by the strong positive correlation between age and taking CVD related medication.

Model 3 was chosen to model GP utilisation due to the high statistical significance of all variables and the greater Pseudo R2 value. The variable covariance matrix from Model 3 is reported in Table S5.

*Table S5: Variance-covariance matrix for GP Attendance, Model 3*

|  | Age | Male | Black | Asian | BMI | Diabetes | Life Limiting Illness | Antidepressants | Lipid Lowering Med | Anti-Hypertensive Med | Constant | LnAlpha |
| --- | --- | --- | --- | --- | --- | --- | --- | --- | --- | --- | --- | --- |
| Age | 0.0000 |  |  |  |  |  |  |  |  |  |  |  |
| Male | -0.0000 | 0.0009 |  |  |  |  |  |  |  |  |  |  |
| Black | 0.0000 | 0.0001 | 0.0081 |  |  |  |  |  |  |  |  |  |
| Asian | 0.0000 | 0.0000 | 0.0003 | 0.0031 |  |  |  |  |  |  |  |  |
| BMI | 0.0000 | -0.0000 | -0.0000 | 0.0000 | 0.0000 |  |  |  |  |  |  |  |
| Diabetes | -0.0000 | -0.0000 | -0.0002 | -0.0001 | -0.0000 | 0.0028 |  |  |  |  |  |  |
| Life Limiting Illness | -0.0000 | 0.0000 | -0.0000 | 0.0000 | -0.0000 | -0.0001 | 0.0010 |  |  |  |  |  |
| Antidepressants | 0.0000 | 0.0001 | 0.0002 | 0.0002 | -0.0000 | -0.0001 | -0.0003 | 0.0017 |  |  |  |  |
| Lipid lowering Medication | -0.0000 | -0.0002 | 0.0001 | -0.0000 | -0.0000 | -0.0005 | -0.0000 | -0.0001 | 0.0018 |  |  |  |
| Anti-Hypertensive Medication | -0.0000 | -0.0000 | -0.0000 | -0.0000 | -0.0000 | -0.0002 | -0.0001 | 0.0000 | -0.0005 | 0.0015 |  |  |
| Constant | -0.0001 | -0.0003 | -0.0005 | -0.0009 | -0.0002 | 0.0004 | 0.0000 | -0.0001 | 0.0006 | 0.0008 | 0.0078 |  |
| LnAlpha | 0.0000 | -0.0000 | -0.0000 | 0.0000 | 0.0000 | 0.0000 | 0.0000 | 0.0000 | 0.0000 | 0.0000 | -0.0000 | 0.0018 |

### 5. Long-term health trajectories

#### 5.1. Longitudinal trajectories for weight

The method uses cross-sectional data from the Health Survey for England 2018 and 2019 to fit distributions that describe changes in weight and BMI from childhood to adulthood. From these distributions it is possible to estimate the expected changes in weight and BMI as individuals age. To maintain heterogeneity in weight throughout an individual’s lifetime, it is important that we assume that the changes in weight are different for different individuals in the model, because we know that some individuals will gain more weight than others over time. With this method, we assume that individual’s weight changes over time, but their position on the weight distribution is constant throughout their life. For example, if an individual has a weight in the 5^th^ percentile aged 20, then their estimated change in weight from age 20 to 21 will be estimated as the difference in weight in the 5^th^ percentile for age 21 and 20.

The advantages of this method are that it uses the most recent health survey for England data on the distributions of BMI across England. We are aware that following the Covid-19 pandemic the health profile and lifestyles of individuals in the UK have changed dramatically, so methods that use retrospective data have limitations. Secondly, this method provides a single method to describe weight changes that can be implemented at any age in the model, rather than methods that aim to combine multiple methods across age categories, which can lead to inconsistencies and increase computation burden. Finally, this method provides a method that can be relatively easily updated with new data from the Health Survey for England, so provides a method that is less likely to become outdated and require further adaptation over time.

There are limitations in this method. The Health Survey for England will capture differences in age categories, but because this data is cross-sectional, rather than longitudinal, will not be able to forecast future changes in BMI based on the distributions found in current ages. For example, we know that in recent years more children are exceeding the thresholds for overweight and obesity. Therefore, it is unlikely that the distribution of weight for those currently aged 50, will be a good representation of the future BMI of today’s children.

#### 5.2. Longitudinal Trajectories of Metabolic Risk Factors

Two separate sets of statistical analyses of longitudinal cohort studies were used to describe metabolic trajectories for individuals in the model. An analysis of the Whitehall II cohort study (6) was developed to describe correlated longitudinal changes in metabolic risk factors for individuals aged 60 years and younger. An analysis of the English Longitudinal Study of Ageing (ELSA) was used to describe trajectories for individuals aged 61 and over. The transition point of 61 years was found to be the age at which there were more data observations for participants in ELSA compared with Whitehall.

##### Whitehall II Data Analysis

Changes in total cholesterol, HDL cholesterol and SBP were estimated from statistical analysis of the Whitehall II cohort. The growth factors for all 5 risk factors were estimated using parallel latent growth modelling. This enabled the growth factors for BMI to be implemented as covariates for the growth processes of glycaemia, systolic blood pressure, and total cholesterol (8). The structural assumptions of the analysis are described in more detail in previous (9).

##### ELSA Data Analysis

Changes in HbA1c, systolic blood pressure, total cholesterol and HDL cholesterol were estimated from a statistical analysis of the ELSA cohort. The changes with age were estimated using independently estimate random coefficient growth models in Stata 13. The growth factors for the metabolic risk factors were assumed to vary between individuals to allow unobservable random effects to describe the heterogeneity in intercept and slope parameters. The structural assumptions of the analysis are described in more detail in previous models (9).

##### HbA1c trajectory in type 2 diagnosed diabetics

Following a diagnosis of diabetes in the simulation all individuals experience an initial fall in HbA1c due to changes in diet and lifestyle as observed in the UKPDS trial (10). We have estimated the expected change in HbA1c conditional on HbA1c at diagnosis by fitting a simple linear regression to three aggregate outcomes reported in the study. These showed that the change in HbA1c increases for higher HbA1c scores at diagnosis. The regression parameters to estimate change in HbA1c are reported in Table S6.

*Table S6: Estimated change in HbA1c following diabetes diagnosis*

|  | Mean | Standard error |
| --- | --- | --- |
| Change in HbA1c Intercept | -2.99 | .048 |
| HbA1c at baseline | 0.55 | 0.40 |

After this initial reduction in HbA1c the longitudinal trajectory of HbA1c is estimated using the UKPDS outcomes model (15) rather than the Whitehall II statistical analysis. The UKPDS dataset is made up of a newly diagnosed diabetic population. As part of the UKPDS Outcomes model, longitudinal trial data were analysed using a random effects model. The coefficients of the model are reported in Table S7.

*Table S7: Coefficient estimates for HbA1c estimated from UKPDS data*

|  | Mean Coefficient | Coefficient standard error |
| --- | --- | --- |
| Intercept | -0.024 | 0.017 |
| Log transformation of year since diagnosis | 0.144 | 0.009 |
| Binary variable for year after diagnosis | -0.333 | 0.05 |
| HbA1c score in last period | 0.759 | 0.004 |
| HbA1c score at diagnosis | 0.085 | 0.004 |

##### Total Cholesterol and HDL Cholesterol Trajectories in Individuals not receiving Statins

At baseline, an individual’s total and HDL cholesterol is determined from the HSE 2014 data. In the simulation, individuals aged 60 years and younger have annual changes in total and HDL cholesterol according to the estimates from the statistical analysis of the Whitehall II cohort. The slope of total and HDL cholesterol are assumed to be linear with time. These growth factors are estimated in the model to be conditional on cholesterol at baseline, age at baseline, sex, and an error parameter to reflect unobservable variability in growth trajectories between individuals. As with latent glycaemia, changes in total cholesterol are also influenced by the trajectory of BMI. For individuals aged 61 and over the ELSA statistical models for Total and HDL cholesterol are used to estimate annual change in cholesterol. As individuals transition between the trajectory models the random coefficient factors are updated allowing current observations to inform the trajectories in Total and HDL cholesterol.

##### Total Cholesterol and HDL Cholesterol Trajectories in Individuals receiving Statins

During the simulation process, individuals are prescribed statins to reduce their risk of cardiovascular disease. It is assumed within the model that the statins are effective in reducing an individual’s total cholesterol, and an average effect is applied to all patients receiving statins. A HTA reviewed the literature on the effectiveness and cost-effectiveness of statins in individuals with acute coronary syndrome (11). This report estimated the change in LDL cholesterol for four statin treatments and doses compared with placebo from a Bayesian meta-analysis. The analysis estimated a reduction in LDL cholesterol of -1.45 for simvastatin. This estimate was used to describe the effect of statins in reducing total cholesterol. It was assumed that the effect was instantaneous upon receiving statins and maintained as long as the individual received statins. It was also assumed that individuals receiving statins no longer experienced annual changes in total cholesterol. HDL cholesterol was assumed constant over time if patients received statins.

Non-adherence to statin treatment is a common problem. A HTA reviewed the literature on continuation and compliance with statin treatment. They concluded that there was a lack of adequate reporting, but that the proportion of patients fully compliant with treatment appears to decrease with time, particularly in the first 12 months after initiating treatment, and can fall below 60% after five years (11). Although a certain amount of non-compliance is included within trial data, clinical trials are not considered to be representative of continuation and compliance in general practice. A yearly reduction in statin compliance used in the HTA analysis is reported in Table S8. It is based on the published estimate of compliance for the first five years of statin treatment for primary prevention in general clinical practice (11) Compliance declines to a minimum of 65% after five years of treatment. It is assumed that there is no further drop after five years.

Table S8 Proportion of patients assumed to be compliant with statin treatment, derived from Table 62 in (11)

| Year after statin initiation | 1 | 2 | 3 | 4 | 5 |
| --- | --- | --- | --- | --- | --- |
| Proportion compliant | 0.8 | 0.7 | 0.68 | 0.65 | 0.65 |

In the simulation, we assume in the base case that only 65% of individuals initiate statins when they are deemed eligible. However, those that initiate statins remain on statins for their lifetime. Those who refuse statins may be prescribed them again at a later date.

##### SBP Trajectories in Individuals Not receiving Anti-hypertensive treatment

At baseline an individual’s SBP is determined from the HSE 2014 data. In the simulation, individuals’ aged 60 and younger experience SBP changes every year according to the estimates from the statistical analysis of the Whitehall II cohort. The annual change in SBP is assumed to be linear with time. The growth factors are estimated in the model to be conditional on SBP at baseline, age at baseline, sex, ethnicity, family history of cardiovascular disease, smoking and an error parameter to reflect unobservable variability in growth trajectories between individuals. From ages 61 onwards the ELSA statistical model for systolic blood pressure is used to estimate annual changes. The random coefficient factors are updated using the bivariate covariance matrix for intercept and slope factors.

##### SBP Trajectories in Individuals receiving anti-hypertensive treatment

During the simulation process, if individuals are identified as having SBP higher than 150mm Hg, or SBP higher than 140mm Hg with age 60+, comorbid diabetes, cardiovascular disease, or 10 year risk of cardiovascular disease greater than 20%, they will be prescribed anti-hypertensive treatment in line with the National Institute for Health and Care Excellence (NICE) guidelines (12). The change in SBP following initiation of calcium channel blockers was estimated in a meta-analysis of anti-hypertensive treatments (13). This study identified an average change in SBP of -8.4 for monotherapy with calcium channel blockers. In the simulation model it is assumed that this reduction in SBP is maintained for as long as the individual receives anti-hypertensive treatment. Once an individual is receiving anti-hypertensive treatment it is assumed that their SBP is stable and does not change over time, which implicitly assumes that patients continue to be well managed for their hypertension. For simplicity we do not explicitly simulate treatment switching. The assumed zero flat trajectory in systolic blood pressure whilst receiving anti-hypertensives is supported by the analysis of the ELSA dataset in which self-reported use of anti-hypertensives was included as a covariate for age-related change in systolic blood pressure. The analysis found that most of the observed changes in systolic blood pressure were removed if individuals were taking anti-hypertensives.

##### Metabolic Risk factor screening

We assume that individuals eligible for anti-hypertensive treatment or statins will be identified through opportunistic screening if they meet certain criteria and attend the GP for at least one visit in the simulation period.

- Individuals with a history of cardiovascular disease;
- Individuals with a major microvascular event (foot ulcer, blindness, renal failure or amputation);
- Individuals with diagnosed diabetes;
- Individuals identified with Impaired Glucose Regulation;
- Individuals with systolic blood pressure greater than 160mmHg.

The base case has been designed to represent a health system with moderate levels of screening for hypertension, and dyslipidaemia. Alternative assumptions for more or less intensive opportunistic screening can be assumed.

##### Diagnosis and Treatment Initiation

It is assumed that there are three, non-mutually exclusive outcomes from the vascular checks or opportunistic screening. Firstly, that the patient receives statins to reduce cardiovascular risk. Secondly, that the patient has high blood pressure and should be treated with anti-hypertensive medication. The following threshold estimates were used to determine these outcomes.

- Statins are initiated if the individual has greater than or equal to 20% 10 year CVD risk estimated from the QRISK2 2012 algorithm.
- Anti-hypertensive treatment is initiated if systolic blood pressure is greater than 150 or if the individual is over 60 years, has a history of CVD, diabetes or a CVD risk >20%, the threshold for systolic blood pressure is 140 (12).

##### Diabetes Diagnosis

#### Diabetes Diagnosis

The HSE repeatedly demonstrates that a large proportion of the English population have a HbA1c test score above 6.5%, and do not report a diagnosis of diabetes. Therefore, it is important to allow some undiagnosed cases of diabetes to persist in the model. Diabetes onset is simulated using the QDiabetes 2018 risk algorithm (14). The model estimates the QDiabetes score, with HBA1c covariates, for undiagnosed individuals within the population in each annual cycle. This score describes the individual's 10 year probability of being diagnosed with diabetes conditional on demographic, socioeconomic, ethnicity, medical history and HbA1c score. The QDiabetes 2018 model was chosen to describe individuals' risk of diabetes in the model to describe differential risks across individuals. The one-year probability of diabetes onset uses the coefficients from the QDiabetes 2018 tool, applied to a baseline risk calibrated to observed data on the rate of diabetes diagnoses in this population.

In each cycle an individual is diagnosed with Diabetes and starts treatment if they have diabetes onset and meet the following criteria.

- Diagnosed hypertension
- Micro/macro vascular diabetes complication
- HbA1c above simulated HbA1c at diagnosis, median 7.5% (15)

The QDiabetes 2018 risk equation can be used to calculate the probability of type 2 diabetes. The equation estimates the probability of diabetes in the next period. The equation for the probability of an event in the next period is calculated as

$$p\left( Y=1 \right)=1-{S(1)}^{\theta}$$

$$\theta=\sum\beta X$$

The probability of an event is calculated from the survival function at 1 year raised to the power of $\theta$, where $\theta$ is the sum product of the coefficients multiplied by the individual’s characteristics. Underlying survival curves for men and women were extracted from the QDiabetes 2018 open source file. Mean estimates for the continuous variables were also reported in the open source files.

#### 5.3. Comorbid Outcomes and Mortality

In every model cycle individuals within the model are evaluated to determine whether they have a clinical event, including mortality, within the cycle period. In each case the simulation estimates the probability that an individual has the event and uses a random number draw to determine whether the event occurred.

##### Cardiovascular Disease

The probability of the first cardiovascular event is estimated from the QRISK2 predicted model of cardiovascular disease (16). The QRISK2 is a validated risk prediction algorithm to identify individuals at high risk of cardiovascular disease. The algorithm was developed from UK data and incorporates social deprivation and ethnicity. The QRISK2 equation estimates the probability of a cardiovascular event in the next 10 years conditional on ethnicity, smoking status, age, BMI, ratio of total/HDL cholesterol, Townsend score, atrial fibrillation, rheumatoid arthritis, renal disease, hypertension, diabetes, and family history of cardiovascular disease. Data on all these variables was available from the HSE 2014.

We estimated the 1 year parameter using a simple calibration process to match the CVD incidence from the WRAP trial after 5 years of follow-up to the simulated incidence of CVD events. A simple calibration approach was taken to identify a parameter estimate for the 1 year survival parameter for men and women that would simulate the estimated CVD incidence reported from the 5 year follow-up data within a 2% margin or error. This data was based on cardiovascular events to include MI, percutaneous coronary intervention (PCI), Experienced bypass (CABG), invasive cardiovascular procedure, Transient Ischaemic Attack or stroke reported in GP records.

At 5 years follow-up GP records for CVD history were available for 859 women, and 408 men. Of those 14 and 18 had experienced a CVD event respectively. For the calibration were identified survival parameters for men and women that would generate an incidence of 3.26 per 1000 person years for women and 8.824 per 1000 person years for men in the brief intervention simulation. These parameters were estimated with a simple iterative process simulating 20,000 randomly selected individuals. The final estimates were tested against other random samples. Starting values were selected from using the survival parameters reported in the QRISK 2012 source code.

Table S9 reports the coefficient estimates for the QRISK2 algorithm. The standard errors were not reported within the open source code. Coefficients that were not reported in this publication were assumed to have standard errors of 20%.

*Table S9: Estimated coefficients for predicting the probability of CVD adjusting for individual characteristics*

|  | Women | | | Men | |  | Women | | Men | | |
| --- | --- | --- | --- | --- | --- | --- | --- | --- | --- | --- | --- |
| Covariates | Mean | Standard error | Mean | | Standard error | Interaction terms | Mean | Standard error | | Mean | Standard error |
| White | 0.0000 | 0.0000 | 0.0000 | | 0.0000 | Age1*smoke1 | 0.6891 | 0.035 | | 0.9244 | 0.776 |
| Indian | 0.2672 | 0.0537 | 0.2785 | | 0.0425 | Age1*smoke2 | 0.6943 | 0.066 | | 1.9598 | 3.341 |
| Pakistani | 0.7148 | 0.0698 | 0.6068 | | 0.0547 | Age1*smoke3 | -1.6952 | 0.231 | | 2.9994 | 3.075 |
| Bangladeshi | 0.3703 | 0.1073 | 0.7104 | | 0.0727 | Age1*smoke4 | -1.2150 | 0.308 | | 5.0371 | 3.529 |
| Other Asian | 0.2074 | 0.1071 | 0.8626 | | 0.0845 | Age1*AF | -3.5855 | 0.922 | | 8.2354 | 1.406 |
| Caribbean | -0.1744 | 0.0619 | 3.8735 | | 0.0641 | Age1*renal disease | -3.0767 | 0.528 | | -3.9747 | 3.403 |
| Black African | -0.3272 | 0.1275 | 0.1347 | | 0.1094 | Age1*hypertension | -4.0295 | 0.450 | | 7.8738 | 6.793 |
| Chinese | -0.2201 | 0.1721 | -0.1558 | | 0.1538 | Age1*Diabetes | -3.3145 | 0.369 | | 5.0624 | 2.558 |
| Other | -0.2090 | 0.0793 | -3.7728 | | 0.0734 | Age1*BMI 1 | -5.5934 | 0.617 | | 33.5438 | 0.654 |
| Non-smoker | 0.0000 | 0.0000 | 0.1526 | | 0.0000 | Age1*BMI 2 | 64.3636 | 0.050 | | -129.9767 | 3.584 |
| Former smoker | 0.1947 | 0.0152 | 0.0132 | | 0.0108 | Age1*family history CVD | 0.8605 | 0.050 | | 1.9280 | 3.584 |
| Light smoker | 0.6229 | 0.0220 | 0.0644 | | 0.0166 | Age1*SBP | -0.0509 | 0.003 | | 0.0523 | 0.030 |
| Moderate smoker | 0.7406 | 0.0178 | 1.4235 | | 0.0148 | Age1*Townsend | 0.1519 | 0.007 | | -0.1731 | 0.510 |
| Heavy smoker | 0.9134 | 0.0194 | 0.3021 | | 0.0143 | Age2*smoke1 | -0.1765 | 0.001 | | -0.0034 | 1.594 |
| Age 1 | 3.8735 |  | -17.6226 | |  | Age2*smoke2 | -0.2324 | 0.000 | | -0.0051 | 4.737 |
| Age 2 | 0.1347 |  | 0.0242 | |  | Age2*smoke3 | 0.2734 | 0.002 | | 0.0003 | 4.627 |
| BMI 1* | -0.1558 | 0.0423 | 1.7320 | | 0.0299 | Age2*smoke4 | 0.1433 | 0.003 | | 0.0031 | 5.373 |
| BMI2 * | -3.7728 |  | -7.2312 | |  | Age2*AF | 0.4987 | 0.010 | | 0.0073 | 2.890 |
| Ratio Total / HDL chol | 0.1526 | 0.0044 | 0.1751 | | 0.0022 | Age2*renal disease | 0.4393 | 0.007 | | -0.0262 | 5.654 |
| SBP | 0.0132 | 0.0045 | 0.0102 | | 0.0046 | Age2*hypertension | 0.6904 | 0.005 | | 0.0086 | 3.763 |
| Townsend | 0.0644 | 0.0068 | 0.0298 | | 0.0048 | Age2*Diabetes | 0.4865 | 0.004 | | -0.0002 | 0.193 |
| AF | 1.4235 | 0.0310 | 0.9891 | | 0.1018 | Age2*BMI 1 | 1.5223 | 0.007 | | 0.0812 | 2.110 |
| Rheumatoid arthritis | 0.3021 | 0.0319 | 0.2542 | | 0.0445 | Age2*BMI 2 | -12.7413 |  | | -0.2559 |  |
| Renal disease | 0.8615 | 0.0639 | 0.7950 | | 0.0702 | Age2*family history CVD | -0.2757 | 0.001 | | -0.0057 | 5.321 |
| Hypertension | 0.5889 | 0.0115 | 0.6229 | | 0.0112 | Age2*SBP | 0.0074 | 0.000 | | -0.0001 | 0.058 |
| Diabetes | 1.1350 | 0.0199 | 0.9373 | | 0.0175 | Age2*Townsend | -0.0487 | 0.000 | | -0.0011 | 0.601 |
| Family history of CVD | 0.5134 | 0.0122 | 0.5923 | | 0.0111 |  |  |  | |  |  |
| AF Atrial Fibrillation CVD Cardiovascular disease SBP systolic blood pressure * covariates transformed with fractional polynomials | | | | | | | | | | | |

The QRISK2 risk equation can be used to calculate the probability of a cardiovascular event including: coronary heart disease (angina or myocardial infarction), stroke, or transient ischaemic attacks, fatality due to cardiovascular disease. The equation estimates the probability of a cardiovascular event in the next period conditional on the coefficients listed in Table S9. The equation for the probability of an event in the next period is calculated as

$$p\left( Y=1 \right)=1-{S(1)}^{\theta}$$

$$\theta=\sum\beta X$$

The probability of an event is calculated from the survival function at 1 year raised to the power of $\theta$, where $\theta$ is the sum product of the coefficients reported in Table S9 multiplied by the individual’s characteristics. Underlying survival curves for men and women were extracted from the QRISK2 open source file. Mean estimates for the continuous variables were also reported in the open source files.

The QRISK2 risk equation can be used to calculate the probability of a cardiovascular event including: coronary heart disease (angina or myocardial infarction), stroke, or transient ischaemic attacks, fatality due to cardiovascular disease. The equation estimates the probability of a cardiovascular event in the next period conditional on the coefficients listed in Table S9. The equation for the probability of an event in the next period is calculated as

$$p\left( Y=1 \right)=1-{S(1)}^{\theta}$$

$$\theta=\sum\beta X$$

The probability of an event is calculated from the survival function at 1 year raised to the power of $\theta$, where $\theta$ is the sum product of the coefficients reported in Table S9 multiplied by the individual’s characteristics. Underlying survival curves for men and women were extracted from the QRISK2 open source file. Mean estimates for the continuous variables were also reported in the open source files.

We modified the QRISK2 assumptions regarding the relationship between IGR, diabetes and cardiovascular disease. Firstly, we assumed that individuals with HbA1c>6.5 have an increased risk of cardiovascular disease even if they have not received a formal diagnosis. Secondly, risk of cardiovascular disease was assumed to increase with HbA1c for test results greater than 6.5 to reflect observations from the UKPDS that HbA1c increases the risk of MI and Stroke (17). Thirdly, prior to type 2 diabetes (HbA1c>6.5) HbA1c is linearly associated with cardiovascular disease. A study from the EPIC Cohort has found that a unit increase in HbA1c increases the risk of coronary heart disease by a hazard ratio of 1.25, after adjustment for other risk factors (18). A linear risk reduction was applied at HbA1c levels below the HSE mean.

The QRISK2 algorithm identifies which individuals experience a cardiovascular event but does not specify the nature of the event. The nature of the cardiovascular event was determined independently. A targeted search of recent Health Technology appraisals of cardiovascular disease was performed to identify a model for the progression of cardiovascular disease following a first event. A Health Technology Assessment (HTA) assessing statins gives age and sex specific distributions of CVD, which were used to assign all QRISK2 events (19). Table S10 reports the probability of cardiovascular outcomes by age and gender.

*Table S10: The probability distribution of cardiovascular events by age and gender*

|  | Age | Stable angina | Unstable angina | MI rate | Fatal CHD | TIA | Stroke | Fatal CVD |
| --- | --- | --- | --- | --- | --- | --- | --- | --- |
| Men | 45-54 | 0.307 | 0.107 | 0.295 | 0.071 | 0.060 | 0.129 | 0.030 |
|  | 55-64 | 0.328 | 0.071 | 0.172 | 0.086 | 0.089 | 0.206 | 0.048 |
|  | 65-74 | 0.214 | 0.083 | 0.173 | 0.097 | 0.100 | 0.270 | 0.063 |
|  | 75-84 | 0.191 | 0.081 | 0.161 | 0.063 | 0.080 | 0.343 | 0.080 |
|  | 85+ | 0.214 | 0.096 | 0.186 | 0.055 | 0.016 | 0.351 | 0.082 |
| Women | 45-54 | 0.325 | 0.117 | 0.080 | 0.037 | 0.160 | 0.229 | 0.054 |
|  | 55-64 | 0.346 | 0.073 | 0.092 | 0.039 | 0.095 | 0.288 | 0.067 |
|  | 65-74 | 0.202 | 0.052 | 0.121 | 0.081 | 0.073 | 0.382 | 0.090 |
|  | 75-84 | 0.149 | 0.034 | 0.102 | 0.043 | 0.098 | 0.464 | 0.109 |
|  | 85+ | 0.136 | 0.029 | 0.100 | 0.030 | 0.087 | 0.501 | 0.117 |

##### Subsequent Cardiovascular events

After an individual has experienced a cardiovascular event, it is not possible to predict the transition to subsequent cardiovascular events using QRISK2. As with assigning first CVD events, the probability of subsequent events was estimated from the HTA evaluating statins (19). This study reported the probability of future events conditional on the nature of the previous event. Table S11 reports an example of the probabilities within a year of transitioning from stable angina, unstable angina, myocardial infarction (MI), transient ischemic attack (TIA) or stroke for individuals by age group.

*Table S11: Probability of cardiovascular event conditional on age and status of previous event (column1)*

|  | Stable angina | Unstable angina 1 | Unstable angina 2 | MI 1 | MI 2 | TIA | Stroke 1 | Stroke 2 | CHD death | CVD death |
| --- | --- | --- | --- | --- | --- | --- | --- | --- | --- | --- |
| Age 45 |  |  |  |  |  |  |  |  |  |  |
| Stable angina | 0.9946 | 0.0013 | 0 | 0.0032 | 0 | 0 | 0 | 0 | 0.0009 | 0 |
| Unstable angina (1st yr) | 0 | 0 | 0.9127 | 0.0495 | 0 | 0 | 0 | 0 | 0.0362 | 0.0016 |
| Unstable angina (subsequent) | 0 | 0 | 0.9729 | 0.0186 | 0 | 0 | 0 | 0 | 0.0081 | 0.0004 |
| MI (1st yr) | 0 | 0 | 0 | 0.128 | 0.8531 | 0 | 0.0015 | 0 | 0.0167 | 0.0007 |
| MI (subsequent) | 0 | 0 | 0 | 0.0162 | 0.978 | 0 | 0.0004 | 0 | 0.0052 | 0.0002 |
| TIA | 0 | 0 | 0 | 0.0016 | 0 | 0.9912 | 0.0035 | 0 | 0.0024 | 0.0013 |
| Stroke (1st yr) | 0 | 0 | 0 | 0.0016 | 0 | 0 | 0.0431 | 0.9461 | 0.0046 | 0.0046 |
| Stroke (subsequent) | 0 | 0 | 0 | 0.0016 | 0 | 0 | 0.0144 | 0.9798 | 0.0021 | 0.0021 |
| Age 55 |  |  |  |  |  |  |  |  |  |  |
| Stable angina | 0.9874 | 0.0029 | 0 | 0.0062 | 0 | 0 | 0 | 0 | 0.0035 | 0 |
| Unstable angina (1st yr) | 0 | 0 | 0.8859 | 0.0497 | 0 | 0 | 0 | 0 | 0.0617 | 0.0027 |
| Unstable angina (subsequent) | 0 | 0 | 0.9548 | 0.0348 | 0 | 0 | 0 | 0 | 0.01 | 0.0004 |
| MI (1st yr) | 0 | 0 | 0 | 0.1152 | 0.8483 | 0 | 0.0032 | 0 | 0.0319 | 0.0014 |
| MI (subsequent) | 0 | 0 | 0 | 0.0179 | 0.9716 | 0 | 0.001 | 0 | 0.0091 | 0.0004 |
| TIA | 0 | 0 | 0 | 0.0031 | 0 | 0.9626 | 0.0181 | 0 | 0.0092 | 0.007 |
| Stroke (1st yr) | 0 | 0 | 0 | 0.0031 | 0 | 0 | 0.0459 | 0.9288 | 0.0111 | 0.0111 |
| Stroke (subsequent) | 0 | 0 | 0 | 0.0031 | 0 | 0 | 0.0186 | 0.9685 | 0.0049 | 0.0049 |
| Age 65 |  |  |  |  |  |  |  |  |  |  |
| Stable angina | 0.976 | 0.006 | 0 | 0.011 | 0 | 0 | 0 | 0 | 0.007 | 0 |
| Unstable angina (1st yr) | 0 | 0 | 0.8435 | 0.0488 | 0 | 0 | 0 | 0 | 0.1031 | 0.0046 |
| Unstable angina (subsequent) | 0 | 0 | 0.9244 | 0.0632 | 0 | 0 | 0 | 0 | 0.0119 | 0.0005 |
| MI (1st yr) | 0 | 0 | 0 | 0.1019 | 0.8287 | 0 | 0.0068 | 0 | 0.0599 | 0.0027 |
| MI (subsequent) | 0 | 0 | 0 | 0.0185 | 0.9634 | 0 | 0.0022 | 0 | 0.0152 | 0.0007 |
| TIA | 0 | 0 | 0 | 0.0055 | 0 | 0.9174 | 0.0423 | 0 | 0.0185 | 0.0163 |
| Stroke (1st yr) | 0 | 0 | 0 | 0.0055 | 0 | 0 | 0.0481 | 0.8944 | 0.026 | 0.026 |
| Stroke (subsequent) | 0 | 0 | 0 | 0.0055 | 0 | 0 | 0.0223 | 0.9514 | 0.0104 | 0.0104 |
| Age 75 |  |  |  |  |  |  |  |  |  |  |
| Stable angina | 0.9681 | 0.0091 | 0 | 0.0158 | 0 | 0 | 0 | 0 | 0.007 | 0 |
| Unstable angina (1st yr) | 0 | 0 | 0.7789 | 0.0466 | 0 | 0 | 0 | 0 | 0.1671 | 0.0074 |
| Unstable angina (subsequent) | 0 | 0 | 0.8733 | 0.1122 | 0 | 0 | 0 | 0 | 0.0139 | 0.0006 |
| MI (1st yr) | 0 | 0 | 0 | 0.0874 | 0.7849 | 0 | 0.0141 | 0 | 0.1088 | 0.0048 |
| MI (subsequent) | 0 | 0 | 0 | 0.0178 | 0.953 | 0 | 0.0047 | 0 | 0.0235 | 0.001 |
| TIA | 0 | 0 | 0 | 0.008 | 0 | 0.8588 | 0.0828 | 0 | 0.0185 | 0.0319 |
| Stroke (1st yr) | 0 | 0 | 0 | 0.008 | 0 | 0 | 0.0446 | 0.8302 | 0.0586 | 0.0586 |
| Stroke (subsequent) | 0 | 0 | 0 | 0.008 | 0 | 0 | 0.0246 | 0.9262 | 0.0206 | 0.0206 |
| Age 85 |  |  |  |  |  |  |  |  |  |  |
| Stable angina | 0.9601 | 0.0122 | 0 | 0.0207 | 0 | 0 | 0 | 0 | 0.007 | 0 |
| Unstable angina (1st yr) | 0 | 0 | 0.6873 | 0.0425 | 0 | 0 | 0 | 0 | 0.2587 | 0.0115 |
| Unstable angina (subsequent) | 0 | 0 | 0.7878 | 0.1955 | 0 | 0 | 0 | 0 | 0.016 | 0.0007 |
| MI (1st yr) | 0 | 0 | 0 | 0.0711 | 0.7053 | 0 | 0.0278 | 0 | 0.1875 | 0.0083 |
| MI (subsequent) | 0 | 0 | 0 | 0.016 | 0.9394 | 0 | 0.0091 | 0 | 0.034 | 0.0015 |
| TIA | 0 | 0 | 0 | 0.0104 | 0 | 0.838 | 0.0961 | 0 | 0.0185 | 0.037 |
| Stroke (1st yr) | 0 | 0 | 0 | 0.0104 | 0 | 0 | 0.0446 | 0.702 | 0.1215 | 0.1215 |
| Stroke (subsequent) | 0 | 0 | 0 | 0.0104 | 0 | 0 | 0.0252 | 0.8894 | 0.0375 | 0.0375 |

##### Congestive Heart Failure

The review of previous economic evaluations of diabetes prevention cost-effectiveness studies found that only a small number of models had included congestive heart failure as a separate outcome. Discussion with the stakeholder group identified that the UKPDS Outcomes model would be an appropriate risk model for congestive heart failure in type 2 diabetes patients. However, it was suggested that this would not be an appropriate risk equation for individuals with normal glucose tolerance or impaired glucose tolerance. The Framingham risk equation was suggested as an alternative. As described above, switching from the Framingham risk score to the UKPDS was not possible due to differences in covariate selection. The main limitations of this equation is that it is quite old, based on a non-UK population, and include diabetes as a discrete health state rather than on a continuous scale.

Congestive heart failure was included as a separate cardiovascular event because it was not included as an outcome of the QRISK2. The Framingham Heart Study has reported logistic regressions to estimate the 4 year probability of congestive heart failure for men and women (20). The equations included age, diabetes diagnosis, BMI and systolic blood pressure to adjust risk based on individual characteristics. We used this risk equation to estimate the probability of congestive heart failure in the SPHR diabetes prevention model. Table S12 describes the covariates for the logit models to estimate the probability of congestive heart failure in men and women.

*Table S12: Logistic regression coefficients to estimate the 4-year probability of congestive heart failure from the Framingham study*

| Variables | Units | Regression  Coefficient | OR (95% CI) | P |
| --- | --- | --- | --- | --- |
| Men | | | | |
| Intercept |  | -9.2087 |  |  |
| Age | 10 y | 0.0412 | 1.51 (1.31-1.74) | <.001 |
| Left ventricular hypertrophy | Yes/no | 0.9026 | 2.47 (1.31-3.77) | <.001 |
| Heart rate | 10 bpm | 0.0166 | 1.18 (1.08-1.29) | <.001 |
| Systolic blood pressure | 20 mm Hg | 0.00804 | 1.17 (1.04-1.32) | 0.007 |
| Congenital heart disease | Yes/no | 1.6079 | 4.99 (3.80-6.55) | <.001 |
| Valve disease | Yes/no | 0.9714 | 2.64 (1.89-3.69) | <.001 |
| Diabetes | Yes/no | 0.2244 | 1.25 (0.89-1.76) | 0.2 |
| Women | | | | |
| Intercept |  | -10.7988 |  |  |
| Age | 10 y | 0.0503 | 1.65 (1.42-1.93) | <.001 |
| left ventricular hypertrophy | Yes/no | 1.3402 | 3.82 (2.50-5.83) | <.001 |
| Heart rate | 100 cL | 0.0105 | 1.11 (1.01-1.23) | 0.03 |
| Systolic blood pressure | 10 bpm | 0.00337 | 1.07 (0.96-1.20) | 0.24 |
| congenital heart disease | 20 mm Hg | 1.5549 | 4.74 (3.49-6.42) | <.001 |
| Valve disease | Yes/no | 1.3929 | 4.03 (2.86-5.67) | <.001 |
| Diabetes | Yes/no | 1.3857 | 4.00 (2.78-5.74) | <.001 |
| BMI | kg/m2 | 0.0578 | 1.06 (1.03-1.09) | <.001 |
| Valve disease and diabetes | Yes/no | -0.986 | 0.37 (0.18-0.78) | 0.009 |
| *OR indicates odds ratio; CI, confidence interval; LVH, left ventricular hypertrophy; CHD, congenital heart disease; and BMI, body mass index. Predicted probability of heart failure can be calculated as: p = 1/(1+exp(-xbeta)), where xbeta = Intercept + Sum (of regression coefficient*value of risk factor) | | | | |

Using the estimated population values we adjusted the intercept values to account for the population risk in men and women. This resulted in a risk equation with age, systolic blood pressure, diabetes (diabetes diagnosis or HbA1c>6.5), and BMI in women to describe the risk of congestive heart failure for the policy analysis model.

##### Microvascular Complications

The UKPDS 2 risk equations are used to estimate the risk and incidence of microvascular diseases (17). This data has the advantage of being estimated from a UK diabetic population. Given that the events described in the UKPDS outcomes model are indicative of late stage microvascular complications, we did not believe it was necessary to seek an alternative model that would be representative of an impaired glucose tolerance population.

We assumed that microvascular complications only occur in individuals with HbA1c>48 mmol/mol (6.5%). Whilst some individuals with hyperglycaemia (HbA1c>42 mmol/mol [6.0%]) may be at risk of developing microvascular complications, it is unlikely that they will progress to renal failure, amputation or blindness before a diagnosis of diabetes. Importantly, we did not assume that only individuals who have a formal diagnosis of diabetes are at risk of these complications. This allows us to incorporate the costs of undetected diabetes into the simulation.

In order to simplify the simulation of neuropathy outcomes we consolidated the models for first amputation with and without prior ulcer into a single equation. The parametric survival models were used to generate estimates of the cumulative hazard in the current and previous period. From which the probability of organ damage being diagnosed was estimated.

|  | $p\left( Death \right)=1-exp(H\left( t \right)-H\left( t-1 \right))$ | (1.1) |
| --- | --- | --- |

The functional form for the microvascular models included exponential and Weibull.

##### Retinopathy

We used the UKPDS outcomes model v2 to estimate the incidence of blindness in individuals with HbA1c>48 mmol/mol (6.5%) (17). The exponential model assumes a baseline hazard $\lambda$, which can be calculated from the model coefficients reported in Table S13 and the individual characteristics for $X$.

$$\lambda=exp\left( \beta_{0}\boldsymbol{+}X\beta_{k} \right)$$

*Table S13: Parameters of the UKPDS2 Exponential Blindness survival model*

|  | Mean coefficient | Standard error | Modified mean coefficient |
| --- | --- | --- | --- |
| Lambda | -11.607 | 0.759 | -10.967 |
| Age at diagnosis | 0.047 | 0.009 | 0.047 |
| HbA1c | 0.171 | 0.032 | 0.171 |
| Heart rate | 0.080 | 0.039 |  |
| SBP | 0.068 | 0.032 | 0.068 |
| White Blood Count | 0.052 | 0.019 |  |
| CHF History | 0.841 | 0.287 | 0.841 |
| IHD History | 0.0610 | 0.208 | 0.061 |
| SBP Systolic Blood Pressure; CHF Congestive Heart Failure; IHD Ischaemic Heart Disease | | | |

The age at diagnosis coefficient was multiplied by age in the current year if the individual had not been diagnosed with diabetes, and by the age at diagnosis if the individual had received a diagnosis.

The expected values for the risk factors not included in the SPHR model (heart rate and white blood count) were taken from Figure 3 of the UKPDS publication in which these are described (17). Assuming these mean values, it was possible to modify the baseline risk without simulating heart rate and white blood cell count.

##### Neuropathy

We used the UKPDS outcomes model v2 to estimate the incidence of ulcer and amputation in individuals with HbA1c>48 mmol/mol (6.5%) (17). The parameters of the ulcer and first amputation models are reported in Table S14.

*Table S14: Parameters of the UKPDS2 Exponential model for Ulcer, Weibull model for first amputation with no prior ulcer and exponential model for 1st amputation with prior ulcer*

|  | Ulcer | | 1st Amputation no prior ulcer | | 1st Amputation prior ulcer | | 2nd Amputation | |
| --- | --- | --- | --- | --- | --- | --- | --- | --- |
|  | Logistic | | Weibull | | Exponential | | Exponential | |
|  | Mean | Standard error | Mean | Standard error | Mean | Standard error | Mean | Standard error |
| Lambda | -11.295 | 1.130 | -14.844 | 1.205 | -0.881 | 1.39 | -3.455 | 0.565 |
| Rho |  |  | 2.067 | 0.193 |  |  |  |  |
| Age at diagnosis | 0.043 | 0.014 | 0.023 | 0.011 | -0.065 | 0.027 |  |  |
| Female | -0.962 | 0.255 | -0.0445 | 0.189 |  |  |  |  |
| Atrial fibrillation |  |  | 1.088 | 0.398 |  |  |  |  |
| BMI | 0.053 | 0.019 |  |  |  |  |  |  |
| HbA1c | 0.160 | 0.056 | 0.248 | 0.042 |  |  | 0.127 | 0.06 |
| HDL |  |  | -0.059 | 0.032 |  |  |  |  |
| Heart rate |  |  | 0.098 | 0.050 |  |  |  |  |
| MMALB |  |  | 0.602 | 0.180 |  |  |  |  |
| PVD | 0.968 | 0.258 | 1.010 | 0.189 | 1.769 | 0.449 |  |  |
| SBP |  |  | 0.086 | 0.043 |  |  |  |  |
| WBC |  |  | 0.040 | 0.017 |  |  |  |  |
| Stroke History |  |  | 1.299 | 0.245 |  |  |  |  |

The exponential model assumes a baseline hazard $\lambda$, which can be calculated from the model coefficients reported in Table S18 and the individual characteristics for $X$.

$$\lambda=exp\left( \beta_{0}+X\beta\right)$$

The Weibull model for amputation assumes a baseline hazard:

$$h\left( t \right)=\rho t^{\rho-1}exp(\lambda)$$

where $\lambda$is also conditional on the coefficients and individual characteristics at time t.

The logistic model for ulcer is described below.

$$PrPr \left( X \right) =\frac{exp(X\beta)}{1+exp(X\beta))}$$

The ulcer and amputation models include a number of covariates that were not included in the simulation. As such it was necessary to adjust the statistical models to account for these measures. We estimated a value for the missing covariates and added the value multiplied by the coefficient to the baseline hazard.

The amputation risk model with a history of ulcer was not included in the simulation, but was used to estimate an additional log hazard ratio to append onto the amputation model without a history of ulcer. The log hazard was estimated for each model assuming the same values for other covariates. The difference in the log hazard between the two models was used to approximate the log hazard ratio for a history of ulcer in the amputation model (10.241). The final model specifications are reported in Table S15.

*Table S15: Coefficients estimates for Ulcer and 1st Amputation*

|  | Ulcer | | 1st Amputation | | 2nd Amputation | |
| --- | --- | --- | --- | --- | --- | --- |
|  | Logistic | | Weibull | | Exponential | |
|  | Mean | Standard error | Mean | Standard error | Mean | Standard error |
| Lambda | -11.276 | 1.13 | -13.954 | 1.205 | -3.455 | 0.565 |
| Rho |  |  | 2.067 | 0.193 |  |  |
| Age at Diagnosis | 0.043 | 0.014 | 0.023 | 0.011 |  |  |
| Female | -0.962 | 0.255 | -0.445 | 0.189 |  |  |
| BMI | 0.053 | 0.019 |  |  |  |  |
| HbA1c | 0.160 | 0056 | 0.248 | 0.042 | 0.127 | 0.06 |
| HDL |  |  | -0.059 | 0.032 |  |  |
| Stroke |  |  | 1.299 | 0.245 |  |  |
| Foot Ulcer |  |  | 10.241 |  |  |  |

##### Nephropathy

We used the UKPDS outcomes model v1 to estimate the incidence of renal failure in individuals with HbA1c>48 mmol/mol (6.5%) (17). Early validation analyses identified that the UKPDS v2 model substantially overestimated the incidence of renal failure in the SPHR model. The Weibull model for renal failure assumes a baseline hazard:

$$h\left( t \right)=\rho t^{\rho-1}exp(\lambda)$$

where $\lambda$is also conditional on the coefficients and individual characteristics at time t. The parameters of the renal failure risk model are reported in Table S16.

*Table S16: Parameters of the UKPDS2 Weibull renal failure survival model*

|  | Mean | Standard error |
| --- | --- | --- |
| Lambda | -10.016 | 0.939 |
| Shape parameter | 1.865 | 0.387 |
| SBP | 0.404 | 0.106 |
| BLIND History | 2.082 | 0.551 |

##### Cancer

The conceptual model identified breast cancer and colorectal cancer risk as being related to BMI. However, these outcomes were not frequently included in previous cost-effectiveness models for diabetes prevention. Discussion with stakeholders identified the EPIC Norfolk epidemiology cohort study as a key source of information about cancer risk in a UK population. Therefore, we searched publications from this cohort to identify studies reporting the incidence of these risks. In order to obtain the best quality evidence for the relationship between BMI and cancer risk we searched for a recent systematic review and meta-analysis using key terms ‘Body Mass Index’ and ‘Cancer’, filtering for meta-analysis studies.

##### Breast cancer

Incidence rates for breast cancer in the UK were estimated from the European Prospective Investigation of Cancer (EPIC) cohort. This is a large multi-centre cohort study looking at diet and cancer. In 2004 the UK incidence of breast cancer by menopausal status was reported in a paper from this study investigating the relationship between body size and breast cancer (21). The estimates of the breast cancer incidence in the UK are reported in Table S17.

*Table S17: UK breast cancer incidence*

|  | Number of Cases | Person Years | Mean BMI | Incidence Rate of per person-year | Standard error | Reference |
| --- | --- | --- | --- | --- | --- | --- |

| UK pre-menopause | 102 | 103114.6 | 24 | 0.00099 | 0.00009 | (21) |
| --- | --- | --- | --- | --- | --- | --- |
| UK post-menopause | 238 | 84214.6 | 24 | 0.00283 | 0.00004 | (21) |

A large meta-analysis that included 221 prospective observational studies has reported relative risks of cancers per unit increase in BMI, including breast cancer by menopausal status (22). We included a risk adjustment in the model so that individuals with higher BMI have a higher probability of pre-and post-menopausal breast cancer. In the simulation we adjusted the probability of breast cancer according to the difference in the individual’s BMI and the average BMI reported in the EPIC cohort. The relative risk and confidence intervals per 5mg/m2 increase in BMI are reported in Table S18.

*Table S18: Relative risk of Breast cancer by BMI*

|  | Mean Relative risk | 2.5th Confidence Interval | 97.5th Confidence Interval | Reference |
| --- | --- | --- | --- | --- |

| UK pre-menopause | 0.89 | 0.84 | 0.94 | (22) |
| --- | --- | --- | --- | --- |
| UK post-menopause | 1.09 | 1.04 | 1.14 | (22) |

##### Colorectal cancer

Incidence rates for colorectal cancer in the UK were reported from the European Prospective Investigation of Cancer (EPIC) cohort. The UK incidence of colorectal cancer is reported by gender in a paper from this study investigating the relationship between body size and colon and rectal cancer (23). The estimates of the colorectal cancer incidence are reported in Table S19

*Table S19: UK colorectal cancer incidence*

|  | Number of Cases | Person Years | Mean Age | Mean BMI | Incidence Rate of per person-year | Standard error | Reference |
| --- | --- | --- | --- | --- | --- | --- | --- |

| Male | 125 | 118468 | 53.1 | 25.4 | 0.00106 | 0.0001 | (23) |
| --- | --- | --- | --- | --- | --- | --- | --- |
| Female | 145 | 277133 | 47.7 | 24.5 | 0.00052 | 0.0002 | (23) |

The risk of colorectal cancer has been linked to obesity. We included a risk adjustment in the model to reflect observations that the incidence of breast cancer is increased in individuals with higher BMI. A large meta-analysis that included 221 prospective observational studies has reported relative risks of BMI and cancers, including colon cancer by gender (22). We selected linear relative risk estimates estimated from pooled European and Australian populations. In the simulation we adjusted the incidence of colorectal cancer by adjusting the probability of colorectal cancer by the difference in the individual’s BMI and the average BMI reported in the EPIC cohort. The relative risk and confidence intervals per 5mg/m^2^ increase in BMI are reported in Table S20.

*Table S20: Relative risk of colon cancer by BMI*

|  | Mean Relative risk | 2.5th Confidence Interval | 97.5th Confidence Interval | Reference |
| --- | --- | --- | --- | --- |

| Male | 1.21 | 1.18 | 1.24 | (22) |
| --- | --- | --- | --- | --- |
| Female | 1.04 | 1.00 | 1.07 | (22) |

#### Osteoarthritis

A study from the Bruneck cohort, a longitudinal study of inhabitants of a town in Italy reported diabetes and BMI as independent risk factors for osteoarthritis (24).

The cohort may not be representative of a UK cohort. However, the individuals are from a European country, the study has a large sample size and has estimated the independent effects of BMI and diabetes on the risk of osteoarthritis. No UK based studies identified in our searches met these requirements. The data used to estimate the incidence of osteoarthritis is reported in Table S21. We did not identify any studies that described diabetes risk on a continuous scale.

*Table S21: Incidence of osteoarthritis and estimated risk factors*

|  | No cases | Person years | Mean BMI | Incidence rate | Standard error | Reference |
| --- | --- | --- | --- | --- | --- | --- |

| No diabetes | 73 | 13835 | 24.8 | 0.0053 | 0.0006 | (24) |
| --- | --- | --- | --- | --- | --- | --- |

|  | Hazard ratio | 2.5th | 97.5th |  |  | Reference |
| --- | --- | --- | --- | --- | --- | --- |

| HR Diabetes | 2.06 | 1.11 | 3.84 |  |  | (24) |
| --- | --- | --- | --- | --- | --- | --- |
| HR BMI | 1.076 | 1.023 | 1.133 |  |  | (24) Personal communication |

#### Depression

Depression was included as a health state in the model. However, the severity of depression was not modelled. Depression is described in the simulation as a chronic state from which individuals do not completely remit. We did not estimate the effect of depression on the longitudinal changes for BMI, glycaemia, SBP and cholesterol. As a consequence, it was not possible to relate the impact of depression to the incidence of diabetes and cardiovascular risk.

In the simulation, individuals can develop depression in any cycle of the model. The baseline incidence of depression among all individuals without a history of depression was estimated from a study examining the bidirectional association between depressive symptoms and type 2 diabetes. Although the study was not from a UK population, the US cohort included ethnically diverse men and women aged 45 to 84 years. We assumed that diagnosis of diabetes and/or CVD increased the incidence of depression in individuals who do not have depression at baseline. We identified a method for inflating risk of depression for individuals with diabetes from the US cohort study described above (25). The risk of depression in individuals who have had a stroke was also inflated according to a US cohort study (26). Odds of depression and odds ratios for inflated risk of depression due to diabetes or stroke are presented in Table S22.

*Table S22: Baseline incidence of depression*

| Baseline Risk of depression | | | |
| --- | --- | --- | --- |
|  | Mean | Standard error |  |
| Depression cases in NGT | 336 |  |  |
| Person years | 9139 |  |  |
| Odds of depression | 0.0382 | 0.002 |  |
| Log odds of depression | -3.266 |  |  |
| Inflated risk for Diabetes | | | |
|  | Mean | 2.5th CI | 97.5th CI |
| Odds ratio of diabetes | 1.52 | 1.09 | 2.12 |
| Log odds ratio of diabetes | 0.419 |  |  |
| Inflate risk of stroke | | | |
| Odds ratio of stroke | 6.3 | 1.7 | 23.2 |
| Log odds ratio stroke | 1.8406 |  |  |
| NGT Normal Glucose Tolerance | | | |

#### Dementia

The risk dementia diagnosis is estimated from risk models estimated from the THIN database (27). The THIN dementia risk score uses data from The Health Improvement Network (THIN) database from across the UK. Routinely collected data was used to predict 5-year risk of recorded diagnosis of Dementia for those aged 60-79 and 80+. The sample size is large and the risk scores are representative of the United Kingdom and diagnosis practices between 2000-2011. There is a relatively short follow-up of patients, the low predictive power of the older risk score, and narrow scope to predict dementia diagnosis but not dementia onset.

The parameters for the THIN 60-79 year old and 80-99 risk models are reported in Table S23.

*Table S23: THIN dementia risk models*

| THIN 60-79 Risk Score | | | THIN 80-99 Risk Score | | |
| --- | --- | --- | --- | --- | --- |
| Parameter label | mean | Standard error | Parameter label | mean | Standard error |
| Baseline hazard | 0.9969 |  | Baseline hazard | -0.9277 |  |
| Age | 0.2092 | 0.0047 | Age | 0.055 | 0.0041 |
| Age^2^ | -0.0034 | 0.0003 | Age^2^ | -0.005 | 0.0010 |
| Female | 0.1285 | 0.0278 | Female | 0.16 | 0.0286 |
| Calendar Year | 0.0448 | 0.0050 | Calendar Year | 0.074 | 0.0056 |
| Townsend quintile 2 | 0.0134 | 0.0390 | BMI | -0.05 | 0.0066 |
| Townsend quintile 3 | 0.1179 | 0.0392 | Anti-hypertensives | -0.249 | 0.0265 |
| Townsend quintile 4 | 0.2018 | 0.0402 | Systolic Blood Pressure | -0.006 | 0.0010 |
| Townsend quintile 5 | 0.2255 | 0.0447 | Lipid ratio | 0.042 | 0.0495 |
| BMI | -0.0616 | 0.0038 | Past Smoker | -0.178 | 0.0281 |
| BMI^2^ | 0.0025 | 0.0003 | Smoker | -0.134 | 0.0485 |
| Anti-hypertensives | -0.1320 | 0.0296 | Alcohol Problems | 0.256 | 0.1352 |
| Past Smoker | -0.0679 | 0.0301 | Diabetes | 0.183 | 0.0413 |
| Smoker | -0.0866 | 0.0415 | Stroke | 0.242 | 0.0332 |
| Alcohol problems | 0.4435 | 0.0799 | Atrial Fibrillation | 0.057 | 0.0383 |
| Diabetes | 0.2867 | 0.0417 | Depression | 0.4 | 0.0332 |
| Depression | 0.8336 | 0.0325 | Anxiety | 0.136 | 0.0520 |
| Stroke | 0.5772 | 0.0394 | NSAIDs use | -0.157 | 0.0408 |
| Atrial Fibrillation | 0.2207 | 0.0514 | Aspirin use | 0.092 | 0.0281 |
| Aspirin use | 0.2528 | 0.0326 |  |  |  |

The risk equations were calibrated to incidence rates from the QResearch dataset (Table S24).

*Table S24: Dementia incidence rates used to derive the adjustment factor*

|  | 5 year crude incidence from QResearch | 5 year risk score (SD) for Health Survey for England population | 5 year simulated incidence (SD) with adjustment factor | Adjustment factor (SE) |
| --- | --- | --- | --- | --- |
| THIN risk model 60-79 | 0.00188 | 0.00255 | 0.00255 | 7.628 (0.104) |
| THIN risk model 80+ | 0.01653 | 0.01523 | 0.01510 | 4.557 (0.020) |

#### Mortality

#### Cardiovascular Mortality

Cardiovascular mortality is included as an event within the QRISK2 (16) and the probability of subsequent cardiovascular events obtained from an HTA assessing statins (19), as described in the Cardiovascular disease section above.

#### Cancer Mortality

Cancer mortality rates were obtained from the Office of National statistics (28). The ONS report one and five year net survival rates for various cancer types, by age group and gender. Net survival was an estimate of the probability of survival from the cancer alone. It can be interpreted as the survival of cancer patients after taking into account the background mortality that the patients would have experienced if they had not had cancer.

The age-adjusted 5-year survival rate for breast cancer and colorectal cancer were used to estimate an annual risk of mortality assuming a constant rate of mortality. We assume that the mortality rate does not increase due to cancer beyond 5 years after cancer diagnosis. The five year survival rate for breast cancer is 84.3%, which translated into a 3.37% annual probability of death from breast cancer. The five year survival rate for persons with colorectal cancer is 55.3%, which translated into a 11.16% annual probability of death from colorectal cancer.

#### Other cause Mortality (including diabetes and Dementia risk)

Other cause mortality describes the risk of death from any cause except CVD, and cancer. All-cause mortality rates by age and sex were extracted from the 2018-2020 Office of National Statistics life tables (29). The mortality statistics report the number of deaths by ICD codes for 5-year age groups. We subtracted the number of cardiovascular disease, diabetes, dementia, breast and colorectal cancer related deaths from the all-cause mortality total to estimate other cause mortality rates by age and sex.

### 6. Direct Costs and Social care costs

Table S28 summarises the costs included in the model. A full description of how these costs are derived is provided in an earlier publication of the model (9).

*Table S25: Summary of healthcare and social care costs input into the model in 2022 prices*

|  | Description | Costs (2022 prices) | Reference |
| --- | --- | --- | --- |

| GP visit | Per visit | £41 | (30) |
| --- | --- | --- | --- |
| Diabetes diagnosis | Per occasion | £16 | (31) |
| Hypertension diagnosis | Per occasion | £48.94 | (32) |
| Diabetes monitoring after diagnosis | Per year | £97.91 | (9) |
| Diabetes monotherapy | Per year | £161.40 | (9) |
| Diabetes dual therapy | Per year | £604.79 | (9) |
| Diabetes insulin therapy | Per year | £1546.80 | (33) (9) |
| Statins | Per year | £19.71 | (34) |
| Hypertension treatment | 1^st^ year after diagnosis | £146.85 | (35) |
| Hypertension | Per year | £97.91 | (35) |

| Stable angina | Per year | £512.92 |
| --- | --- | --- |

| Unstable angina year 1 | Per year | £2545.31 | (36) |
| --- | --- | --- | --- |
| MI year 1 | Per year | £5056.65 | (36) |
| Subsequent care cost | Per year | £418.34 | (36) |
| Stroke year 1 | Per year | £15,200.58 | (37) |
| Stroke subsequent | Per year | £1,262.67 | (37) |
| Transient Ischemic Attack | Per year | £1902.33 | (36) |
| Fatal CHD | Per occasion | £2436.02 | (38) |
| Fatal CVD | Per occasion | £2043.15 | (38) |
| Heart failure year 1 | Per year | £1993.96 | (39) |
| Heart failure subsequent | Per year | £12829.02 | (39) |
| Renal Failure | 1 year after diagnosis | £21,465 | (40) |
| Renal Failure | Per year | £8,559 | (40) |
| Foot ulcers | Per year | £3,991 | (33) |
| Amputation year 1 | Per year | £14,162 | (41) |
| Amputation subsequent | Per year | £3,936 | (41) |
| Blindness year 1 | Per year | £3,637 | (41) |
| Blindness subsequent | Per year | £1,378 | (41) |
| Breast cancer | Per diagnosis | £26,967 | (42) |
| Colorectal cancer | Per diagnosis | £19,211 | (42) |
| Osteoarthritis | Per year | £908 | (43) |
| Depression | Per year | £625 | (44) |
| Dementia diagnosis | Per occasion | £752 | (45) |
| Dementia healthcare costs mild | Per year | £3,391.13 | (45) |
| Dementia healthcare costs moderate | Per year | £9,064.21 | (45) |
| Dementia healthcare costs severe | Per year | £10,756.31 | (45) |

### 7. Social Care costs

In this analysis the social care costs account for the costs of formal and informal home help, such as home care workers and sheltered warden managers, or family and friends. Whether an individuals received social care is dependent on their BMI and demographics. Additional social care costs, such as residential care or occupational therapy that are associated with osteoarthritis and stroke are also accounted for from the point these events occur. Additional social care costs associated with the other health outcomes of the model are not included in this estimate. However, reliable social care costs for other conditions are very hard to obtain because they are less commonly incurred in the prevalent patient population and more likely to be attributed to other factors or ageing more generally.

#### Social Care Model Conditional on BMI

To simulate social care utilisation and costs, a cross-sectional logistic regression analysis relating social care use to BMI was reproduced from a previous analysis with updated data (46). This was reproduced with HSE 2019 data to allow the regression outputs to be simulated in the model. A logistic regression was performed to determine the association between whether an individual received social care, formal and/or informal, and BMI, age, sex, deprivation, and ethnicity. A limiting illness indicator was not included in the regression models due to concerns of collinearity. Three regression specifications were assessed and the model with the best model fit was chosen (Table S55). Regression specification 3 was then performed with multiple imputation, to accommodate missing data, using multiple imputation by chained equations (MICE) in R using the “mice” package. The regression coefficients were used to find a fitted value for each individual based on their characteristics, and this was converted to a probability using the equations below. This probability was compared to a random number to determine if they receive social care.

$$P= \frac{1}{1+exp(-(Fitted Value)}$$

Where,

$$Fitted Value= \beta_{0}+\beta_{1}BMI+\beta_{1}BMI+\beta_{2}{BMI}^{2}+\beta_{3}{BMI}^{3}+\beta_{4}AGE 70 to 74+\beta_{5}AGE 75 to 79+\beta_{6}AGE 80 to 84+\beta_{7}AGE 85 over+ e$$

Samples of each coefficient (Table S26) were sampled from a multivariate normal distribution using the variance-covariance from regression specification 3 (Table S27).

*Table S26: Regression outputs associated with social care utilisation across 3 model specifications*

|  | Model 1 | | | Model 2 | | | Model 3 | | | Model 3 MI | | |
| --- | --- | --- | --- | --- | --- | --- | --- | --- | --- | --- | --- | --- |
| Variable | Coefficients | SE | p-value | Coefficient | SE | p-value | Coefficient | SE | p-value | coefficient | SE | p-value |
| Intercept | 5.259 | 1.8245 | ** | 3.2079 | 1.8445 |  | 1.8758 | 1.8542 |  | 2.4908 |  |  |
| BMI | -0.7384 | 0.1667 | *** | -0.605 | 0.1673 | *** | -0.5203 | 0.1678 | ** | -0.5334 | 0.1712 |  |
| BMI2 | 0.0216 | 0.0049 | *** | 0.0182 | 0.0049 | *** | 0.0156 | 0.0049 | ** | 0.016 | 0.0051 | ** |
| BMI3 | -0.0002 | 0 | *** | -0.0002 | 0 | ** | -0.0001 | 0 | ** | -0.0001 | 0 | ** |
| Female (Male = base) |  |  |  | 0.6739 | 0.0837 | *** | 0.6938 | 0.0846 | *** | 0.6116 | 0.0672 | *** |
| 70-74 years old (60-65 = base) | 0.054 | 0.1173 |  | 0.0621 | 0.1182 |  | 0.0644 | 0.1191 |  | 0.1047 | 0.0986 |  |
| 75-79 years old | 0.5371 | 0.1204 | *** | 0.5371 | 0.1216 | *** | 0.5624 | 0.1228 | *** | 0.6443 | 0.1006 | *** |
| 80-84 years old | 1.1228 | 0.1271 | *** | 1.1691 | 0.1287 | *** | 1.1902 | 0.1302 | *** | 1.2405 | 0.1058 | *** |
| 85-89 years old | 1.6107 | 0.1457 | *** | 1.6323 | 0.1477 | *** | 1.6505 | 0.1499 | *** | 1.9247 | 0.1192 | *** |
| 90+ years old | 2.4244 | 0.2136 | *** | 2.4505 | 0.2171 | *** | 2.5115 | 0.2205 | *** | 2.7318 | 0.1676 | *** |
| Black |  |  |  | 0.3154 | 0.3419 |  | -0.0034 | 0.3426 |  | 0.2336 | 0.2586 |  |
| Asian |  |  |  | 0.6371 | 0.2273 | ** | 0.5177 | 0.23 | * | 0.4974 | 0.1849 | ** |
| IMD score 2 |  |  |  |  |  |  | 0.2481 | 0.1305 |  | 0.0982 | 0.1034 |  |
| IMD score 3 |  |  |  |  |  |  | 0.5183 | 0.1297 | *** | 0.3816 | 0.1026 | *** |
| IMD score 4 |  |  |  |  |  |  | 0.7385 | 0.1332 | *** | 0.6773 | 0.1039 | *** |
| IMD score 5 |  |  |  |  |  |  | 1.1371 | 0.1348 | *** | 1.0854 | 0.1066 | *** |
| HSE Year 2018 | -0.0171 | 0.0977 |  | -0.00778 | 0.098581 |  | -0.0226 | 0.0997 |  | -0.0887 | 0.0797 |  |
| HSE Year 2019 | -0.0011 | 0.0972 |  | 0.012318 | 0.098151 |  | -0.0189 | 0.0995 |  | -0.076 | 0.0794 |  |
| obs | 5276 |  |  | 5270 |  |  | 5270 |  |  |  |  |  |
| R2 | 0.075 |  |  | 0.092 |  |  | 0.111 |  |  |  |  |  |
| AIC | 4210 |  |  | 4136 |  |  | 4058 |  |  |  |  |  |

*Table S27: Variance – Covariance matrix of regression specification 3.*

| Part A | Intercept | BMI | BMI2 | BMI3 | Female | 70-74 years | 75-79 years | 80-84 years | 85-89 years | 90+ years |
| --- | --- | --- | --- | --- | --- | --- | --- | --- | --- | --- |
| Intercept | 3.3964 | -0.3122 | 0.0091 | -0.0001 | -0.0127 | 0.0016 | 0.0091 | 0.0092 | 0.0143 | 0.0121 |
| BMI | -0.3122 | 0.0293 | -0.0009 | 0.0000 | 0.0008 | -0.0006 | -0.0014 | -0.0015 | -0.0021 | -0.0021 |
| BMI2 | 0.0091 | -0.0009 | 0.0000 | 0.0000 | 0.0000 | 0.0000 | 0.0000 | 0.0000 | 0.0001 | 0.0001 |
| BMI3 | -0.0001 | 0.0000 | 0.0000 | 0.0000 | 0.0000 | 0.0000 | 0.0000 | 0.0000 | 0.0000 | 0.0000 |
| Female | -0.0127 | 0.0008 | 0.0000 | 0.0000 | 0.0045 | 0.0000 | 0.0000 | 0.0002 | 0.0002 | 0.0005 |
| 70-74 years | 0.0016 | -0.0006 | 0.0000 | 0.0000 | 0.0000 | 0.0097 | 0.0050 | 0.0051 | 0.0051 | 0.0051 |
| 75-79 years | 0.0091 | -0.0014 | 0.0000 | 0.0000 | 0.0000 | 0.0050 | 0.0101 | 0.0052 | 0.0052 | 0.0054 |
| 80-84 years | 0.0092 | -0.0015 | 0.0000 | 0.0000 | 0.0002 | 0.0051 | 0.0052 | 0.0112 | 0.0054 | 0.0056 |
| 85-89 years | 0.0143 | -0.0021 | 0.0001 | 0.0000 | 0.0002 | 0.0051 | 0.0052 | 0.0054 | 0.0142 | 0.0058 |
| 90+ years | 0.0121 | -0.0021 | 0.0001 | 0.0000 | 0.0005 | 0.0051 | 0.0054 | 0.0056 | 0.0058 | 0.0281 |
| Black | 0.0022 | 0.0000 | 0.0000 | 0.0000 | -0.0001 | 0.0005 | -0.0002 | -0.0002 | -0.0001 | 0.0005 |
| Asian | 0.0031 | -0.0006 | 0.0000 | 0.0000 | 0.0002 | 0.0010 | 0.0011 | 0.0016 | 0.0013 | 0.0019 |
| IMD 2 | -0.0032 | -0.0003 | 0.0000 | 0.0000 | 0.0001 | 0.0001 | 0.0003 | 0.0003 | 0.0004 | 0.0003 |
| IMD 3 | -0.0013 | -0.0003 | 0.0000 | 0.0000 | 0.0001 | 0.0000 | 0.0001 | 0.0002 | 0.0002 | 0.0002 |
| IMD 4 | -0.0057 | 0.0001 | 0.0000 | 0.0000 | 0.0001 | 0.0000 | 0.0002 | 0.0003 | 0.0005 | 0.0007 |
| IMD 5 | -0.0123 | 0.0006 | 0.0000 | 0.0000 | 0.0002 | 0.0001 | 0.0004 | 0.0005 | 0.0008 | 0.0012 |
| 2018 | -0.0054 | 0.0002 | 0.0000 | 0.0000 | 0.0000 | 0.0001 | 0.0001 | 0.0001 | 0.0001 | 0.0000 |
| 2019 | -0.0044 | 0.0001 | 0.0000 | 0.0000 | 0.0000 | 0.0000 | 0.0000 | 0.0000 | 0.0000 | 0.0000 |

| Part B | Black | Asian | IMD 2 | IMD 3 | IMD 4 | IMD 5 | 2018 | 2019 |
| --- | --- | --- | --- | --- | --- | --- | --- | --- |
| Intercept | 0.0022 | 0.0031 | -0.0032 | -0.0013 | -0.0057 | -0.0123 | -0.0054 | -0.0044 |
| BMI | 0.0000 | -0.0006 | -0.0003 | -0.0003 | 0.0001 | 0.0006 | 0.0002 | 0.0001 |
| BMI2 | 0.0000 | 0.0000 | 0.0000 | 0.0000 | 0.0000 | 0.0000 | 0.0000 | 0.0000 |
| BMI3 | 0.0000 | 0.0000 | 0.0000 | 0.0000 | 0.0000 | 0.0000 | 0.0000 | 0.0000 |
| Female | -0.0001 | 0.0002 | 0.0001 | 0.0001 | 0.0001 | 0.0002 | 0.0000 | 0.0000 |
| 70-74 years | 0.0005 | 0.0010 | 0.0001 | 0.0000 | 0.0000 | 0.0001 | 0.0001 | 0.0000 |
| 75-79 years | -0.0002 | 0.0011 | 0.0003 | 0.0001 | 0.0002 | 0.0004 | 0.0001 | 0.0000 |
| 80-84 years | -0.0002 | 0.0016 | 0.0003 | 0.0002 | 0.0003 | 0.0005 | 0.0001 | 0.0000 |
| 85-89 years | -0.0001 | 0.0013 | 0.0004 | 0.0002 | 0.0005 | 0.0008 | 0.0001 | 0.0000 |
| 90+ years | 0.0005 | 0.0019 | 0.0003 | 0.0002 | 0.0007 | 0.0012 | 0.0000 | 0.0000 |
| Black | 0.0669 | 0.0013 | -0.0001 | -0.0004 | -0.0014 | -0.0027 | -0.0002 | -0.0001 |
| Asian | 0.0013 | 0.0342 | 0.0002 | -0.0005 | -0.0010 | -0.0004 | -0.0001 | 0.0001 |
| IMD 2 | -0.0001 | 0.0002 | 0.0107 | 0.0055 | 0.0055 | 0.0055 | -0.0002 | 0.0001 |
| IMD 3 | -0.0004 | -0.0005 | 0.0055 | 0.0105 | 0.0055 | 0.0056 | -0.0003 | 0.0000 |
| IMD 4 | -0.0014 | -0.0010 | 0.0055 | 0.0055 | 0.0108 | 0.0057 | -0.0001 | 0.0001 |
| IMD 5 | -0.0027 | -0.0004 | 0.0055 | 0.0056 | 0.0057 | 0.0114 | -0.0003 | -0.0003 |
| 2018 | -0.0002 | -0.0001 | -0.0002 | -0.0003 | -0.0001 | -0.0003 | 0.0064 | 0.0031 |
| 2019 | -0.0001 | 0.0001 | 0.0001 | 0.0000 | 0.0001 | -0.0003 | 0.0031 | 0.0063 |

The hourly cost of formal care was assumed to be the hourly weekday cost of a home care worker at £32, sampled using a gamma distribution with a standard error of 3.2 (47). To cost informal health care an opportunity cost approach was taken. The hourly cost of informal health care was assumed to be the average hourly wage of £17.74.

#### Social care cost of Stroke and Dementia

The community costs in the first year following stroke were estimated in a cost-effectiveness analysis using the Sentinel Stroke National Audit Program (48). They report the social care cost associated with residential care. The 5-year cost of residential home care for all stroke types was reported as £10,992. This was assumed to spread evenly over the 5 years and inflated to 2022 prices, resulting in an annual cost used of £2,483. This cost was assumed to have a standard error of 1/10th of the mean and sampled using a gamma distribution.

The social care costs of dementia were estimated in an Alzheimer’s UK report in 2014 (45). The costs are reported in Table S28.

*Table S28: Average annual dementia costs*

|  | Social care costs | Proportion of patients residential care | Total cost 2012 prices | Total Costs 2022 prices |
| --- | --- | --- | --- | --- |
|  | Residential |  |  |  |
| Mild (MMSE 21-26) | 24,737 | 10.4% | £5362 | £6109.46 |
| Moderate (MMSE 10-20) | 25,715 | 76.2% | £21455 | £24445.83 |
| Severe (MMSE 0-9) | 25,874 | 76.2% | £22176 | £25267.34 |

### 8. Utilities

#### Baseline Utility

Baseline utilities for all individuals in the cohort were extracted from the HSE 2011. The tariffs for the responses to the 3 level EQ-5D were derived from a UK population study (49). Utility was assumed to decline due to ageing independent of health status. In the simulation, utility declines by an absolute decrement of 0.004 per year. This estimate is based on previous HTA modelling in cardiovascular disease (50).

#### Utility Decrements

The utility decrements for long term chronic conditions were applied to the age adjusted EQ-5D score. In consultation with stakeholders, we assumed that a diagnosis of diabetes was not associated with a reduction in EQ-5D independent of the utility decrements associated with complications, comorbidities or depression. Cardiovascular disease, renal failure, amputation, foot ulcers, blindness, cancer, osteoarthritis and depression were all assumed to result in utility decrements. The utility decrements are measured as a factor which is applied to the individual’s age adjusted baseline. If individuals have multiple chronic conditions the utility decrements are multiplied together to give the individual’s overall utility decrement from comorbidities and complications, in line with current NICE guidelines for combining comorbidities (51).

Due to the number of health states it was not practical to conduct a systematic review to identify utility decrements for all health states. A pragmatic approach was taken to search for health states within existing health technology assessments for the relevant disease area or by considering studies used in previous economic models for diabetes prevention. Discussions with experts in health economic modeling were also used to identify prominent sources of data for health state utilities.

Two sources of data were identified for diabetes related complications. A study from the UKPDS estimated the impact of changes in health states from a longitudinal cohort (52). They estimated the impact of myocardial infarction, ischaemic heart disease, stroke, heart failure, amputation and blindness on quality of life using seven rounds of EQ-5D questionnaires administered between 1997 and 2007. This data was used to estimate the utility decrement for amputation and congestive heart failure. The absolute decrement for amputation was converted into utility decrement factors that could be multiplied by the individuals’ current EQ-5D to estimate the relative effect of the complication. Blindness was included in the statistical model used for this analysis however the UKPDS analysis reported an increase in health state utility following a diagnosis with blindness. Discussions with the authors highlighted that this was due to treatment following formal classification with blindness and it was decided that this increase in health state utility should not be included in the cost-effectiveness model.

Utility decrements for renal failure and foot ulcers were not available from the UKPDS study described above. A study by Coffey et al. (2000) was used to estimate utility decrements for renal failure and foot ulcers (53). In this study, 2,048 subjects with type 1 and type 2 diabetes were recruited from specialty clinics. The Self-Administered Quality of Well Being index (QWB-SA) was used to calculate a health utility score.

A meta-analysis of utility values for diabetes and diabetes related complications estimated utility decrements for amputation, ulcer, end stage renal failure and blindness (54). The study pooled utility measures using different health state valuation measures in a meta-analysis. Pooling health state utility values is problematic because of the fact that different valuation methods and different preference-based measures (PBMs) can generate different values on exactly the same clinical health state (55).There were not sufficient studies in the meta-analysis to adjust for the effects of health state valuation measure on the result. This is a limitation of the analysis and we decided that it was preferable to use estimates from single studies.

Utility decrements for cardiovascular events were taken from an HTA assessing statins to reflect the utility decrements in all patients (7) rather than using the UKPDS, which is only representative of a diabetic population. The study conducted a literature review to identify appropriate utility multipliers for stable angina, unstable angina, myocardial infarction and stoke. We used these estimates in the model and assume that transient ischaemic attack is not associated with a utility decrement in line with this HTA.

We identified a systematic review of breast cancer utility studies following consultation with colleagues with experience in this area. The review highlighted a single burden of illness study with a broad utility decrement for cancer (56), rather than utilities by cancer type or disease status. This study was most compatible with the structure of the cost-effectiveness structure. Within this study 1823 cancer survivors and 5469 age-, sex-, and educational attainment-matched control subjects completed EQ-5D questionnaires to estimate utility with and without cancer.

The utility decrement for osteoarthritis was taken from a Health Technology Assessment that assessed the clinical effectiveness and cost-effectiveness of glucosamine sulphate/hydrochloride and chondroitin sulphate in modifying the progression of osteoarthritis of the knee (57).

A review of cost-effectiveness studies highlights the scarcity of studies of health-related quality of life in depression (58). The utility studies identified in the review described depression states by severity and did not adjust for comorbid conditions. Furthermore, the valuations were variable between studies suggesting poor consistency in the estimations. Therefore, it was difficult to apply these in the model. We decided to use a study which had used the EQ-5D in an RCT, for consistency with our utility measure (59). They report an average post treatment utility of 0.67, from which we estimated the utility decrement compared with the average utility reported in the HSE dataset. The decrement was then converted into a relative utility reduction.

The quality of life impact of dementia is estimated from a study by Jonsson and colleagues (60). These utility values were identified and used in a NICE HTA for Alzheimer disease (61). A systematic review of health state utilities for Alzheimer’s disease discusses differences in health related quality of life in different settings (62). It is often assumed that patients in institutional settings will be more disabled and have poorer quality of life. However, the studies that compared utility between settings did not identify a statistically significant difference. Therefore, we only related utility to MMSE.

Table S31 reports the multiplicative utility factors that are used in the model to describe health utility decrements from comorbid complications. The mean absolute decrement estimated in each study is reported alongside the baseline utility for each study. The utility factor was estimated by dividing the implied health utility with the comorbidity by the baseline utility.

*Table S29: Utility decrement factors*

|  | Mean Absolute decrement | St. error absolute decrement | Baseline Utility | Multiplicative Utility Factor | Source |
| --- | --- | --- | --- | --- | --- |

| Foot ulcer | -0.099 | 0.013 | 0.689 | 0.856 | Coffey (53) |
| --- | --- | --- | --- | --- | --- |
| Amputation | -0.172 | 0.045 | 0.807 | 0.787 | UKPDS (52) |

| Blind |  |  |  | 1.00 | Assumption |
| --- | --- | --- | --- | --- | --- |

| Renal failure | -0.078 | 0.026 | 0.689 | 0.887 | Coffey (53) |
| --- | --- | --- | --- | --- | --- |
| Stable Angina |  |  |  | 0.801 | Ward HTA (50) |
| Unstable Angina y1 |  |  |  | 0.770 | Ward HTA (50) |
| Unstable Angina y2 |  |  |  | 0.770 | Ward HTA (50) |
| Myocardial Infarction y1 |  |  |  | 0.760 | Ward HTA (50) |
| Myocardial Infarction y2 |  |  |  | 0.760 | Ward HTA (50) |
| Transient Ischaemic Attack |  |  |  | 1.000 | Ward HTA (50) |
| Stroke y1 |  |  |  | 0.629 | Ward HTA (50) |
| Stroke y2 |  |  |  | 0.629 | Ward HTA (50) |
| Breast Cancer | -0.060 | 0.008 | 0.791 | 0.913 | Yabroff (56) |
| Colorectal Cancer | -0.060 | 0.008 | 0.791 | 0.913 | Yabroff (56) |
| Osteoarthritis | -0.101 | 0.069 | 0.791 |  | Black HTA (57) |
| Depression | -0.116 |  | 0.791 | 0.875 | Benedict (59) |
| Congestive Heart Failure | -0.101 | 0.032 |  | 0.875 | UKPDS (52) |
| MMSE 26-30 |  |  | 0.690 |  | Jonsson (60) |
| MMSE 21-25 | -0.05 |  | 0.690 | 0.93 | Jonsson (60) |
| MMSE 15-20 | -0.19 |  | 0.690 | 0.725 | Jonsson (60) |
| MMSE 10-14 | -0.20 |  | 0.690 | 0.710 | Jonsson (60) |
| MMSE 0-9 | -0.36 |  | 0.690 | 0.478 | Jonsson (60) |

UKPDS baseline utility 0.807; HSE baseline 0.7905

### 9. Treatment effect

The effect of calorie labelling is implemented in the model through consumer behaviour change and reformulation. Consumer behaviour change is assumed to be temporary and last only 6 months. Reformulation is assumed to be permanent.

#### Consumer Behaviour change

Estimates are based on Kalbus et al. 2025 where an interrupted time series analysis was conducted using consumer out-of-home purchase data (Kantar’s Worldpanel OOH Purchase panel, 47w/e, 27th Nov 2022) to estimate the immediate (intercept) and longer-term (time trend) impact of England’s calorie labelling policy on calories purchased (Table S30). Based on the observed coefficients we assume that the effect diminishes linearly over the initial six months and plateaus thereafter, the total cumulative difference in calories during this period was calculated by integrating the estimated linear trend. This total difference was then annualized and applied as a daily change in calorie intake (kcal) within the first year cycle of the model to represent the policy's initial impact. For the probabilistic sensitivity analysis these parameters were sampled from a multivariate normal distribution using the covariance matrix generated from the regression output.

*Table S30: Intercept and time trend coefficients*

|  | Level change (95%CI) | Trend change (95%CI) |
| --- | --- | --- |
| All calories purchased OOH – total population | -95.6 (-471.2 to 280.0) | 5.1(-5.5 to 15.8) |
| SA3: All calories purchased OOH – male low SES | -63.7 (-676.6 to 549.2) | 9.2 (-8.2 to 26.5) |
| SA3: All calories purchased OOH – male high SES | 63.61 (-568.4 to 695.6 | 2.1 (-15.8 to 20.0) |
| SA3: All calories purchased OOH – female low SES | 37.0 (-622.4 to 696.5) | 5.7 (-12.9 to 24.4) |
| SA3: All calories purchased OOH – female high SES | -365.8 (-980.2 to 248.7) | 2.46 (-14.9 to 19.9) |

Source: Kalbus (63) sub-group analyses by SES and sex not included in manuscript.

#### Reformulation

The real-world evidence for reformulation following implementation of calorie labelling in the out of home sector was obtained from a pre-post implementation analysis of chain food menus using MenuTracker data. In this study the mean reduction in calories was estimated at 9 kcal per menu item. To convert this estimate to daily calorie reduction we multiplied the estimate by the average number of dishes purchased per week, and divided it by seven.

A sensitivity analysis examined average calorie content on menus of two online food delivery services which estimated a larger overall reduction in calories on menus. The analysis is currently unpublished (Table S31). The average calorie content of menu items decreased more in more deprived areas. Mean change in average calories per menu item (food and drink) from June 2022 to June 2023 per restaurant delivering to LSOA by IMD quintile was calculated based on 27,980 LSOAs in England which had online food delivery available in both time points. A negative figure indicates a drop in mean calories between the two time points.

*Table S31: Estimated change in average calories from pre- post implementation analysis of menu offering*

|  | Mean | 95% Confidence Interval | Source |
| --- | --- | --- | --- |
| Overall change in calories on menu | -9 kcal | (-1, -16) | Essman et al (64) |
| Overall times per week purchase occasions per week from out-of-home sector | 1.56 | NA | Author’s own analysis * |
| SA 3: Male and low SES purchase occasions per week from out-of-home sector | 1.50 | NA | Author’s own analysis * |
| SA 3: Male and high SES purchase occasions per week from out-of-home sector | 1.65 | NA | Author’s own analysis* |
| SA 3: Female and low SES purchase occasions per week from out-of-home sector | 1.43 | NA | Author’s own analysis* |
| SA 3: Female and high SES purchase occasions per week from out-of-home sector | 1.57 | NA | Author’s own analysis* |
| SA 4: IMD1 (least deprived) | -16.23 | -17.86 to -14.60 | Unpublished analysis (Kalbus et al. (2025) Calorie labelling and changes in the calorie content of food and drink available for online food delivery over time) |
| SA 4: IMD2 | -17.13 | -18.81 to -15.45 |  |
| SA 4: IMD3 | -16.25 | -17.82 to -14.67 |  |
| SA 4: IMD4 | -19.07 | -20.43 to -17.70 |  |
| SA 4: IMD5 (most deprived) | -20.22 | -21.44 to -18.99 |  |

*Authors’ own analysis Kantar’s Worldpanel OOH Purchase panel, 47w/e, 27th Nov 2022

Table S32 summarises the intervention effects in base case sensitivity analysis

*Table S32: Key input parameters for effectiveness and cost of the calorie labelling policy in England*

|  | Population | Average effect | Source |
| --- | --- | --- | --- |
| SA1: Consumer response: Change in total calories (kcal) per day | Overall | -2.05 kcal | Kalbus et al (63) |
| SA2: Reformulation: Change in calories per day | Overall | -2.00 kcal | Essmann et al (64) & Author’s own analysis of Kantar’s Worldpanel OOH Purchase panel, 47w/e, 27th Nov 2022 |
| SA3: Consumer response: Change in total calories (kcal) per day | Male and low SES | 3.95 kcal | Kalbus et al (63) |
|  | Male and high SES | 6.5 kcal | Kalbus et al (63) |
|  | Female and low SES | 7.93 kcal | Kalbus et al (63) |
|  | Female and high SES | -23.8 kcal | Kalbus et al (63) |
| SA3: Reformulation: Change in calories per day | Male and low SES | -1.93 kcal | Kalbus et al (63) |
|  | Male and high SES | -2.12 kcal | Kalbus et al (63) |
|  | Female and low SES | -1.84 kcal | Kalbus et al (63) |
|  | Female and high SES | -2.02 kcal | Kalbus et al (63) |
| SA4: Reformulation: Change in calories per week | IMD1 (least deprived) | -16.23 | Unpublished Kalbus et al. (2025) |
|  | IMD2 | -17.13 | Unpublished Kalbus et al. (2025) |
|  | IMD3 | -16.25 | Unpublished Kalbus et al. (2025) |
|  | IMD4 | -19.07 | Unpublished Kalbus et al. (2025) |
|  | IMD5 (most deprived) | -20.22 | Unpublished Kalbus et al. (2025) |
| SA7: Consumer response: Change in total calories (kcal) per dish | Overall | -47 kcal (95% CI 15–78) | Crockett et al. (65) |
| SA7: Reformulation: Change in calories per dish | Overall | -15 kcal (95% CI 8–23) | Zlatevska et al. (66) |

| Population Intervention costs | Implementation cost (£) in year 1 | £25,000 | Impact Assessment (67) |
| --- | --- | --- | --- |
|  | Enforcement cost (£) per year | £100,000 | Impact Assessment (67) |

|  | Total General Population aged 13-79 (N) | 45,198,512 | 2021 Census: England estimates |
| --- | --- | --- | --- |

### 10. Model Stability

Figure S2 illustrates the stability of cumulative incidence of type 2 diabetes, incremental costs, incremental QALYs and incremental next benefit in the base case. In the final base case analysis we simulate 200,000 individuals, sensitivity analyses were simulated for 100,000 individuals.

*Figure S2: Model stability test across number of simulated individuals*

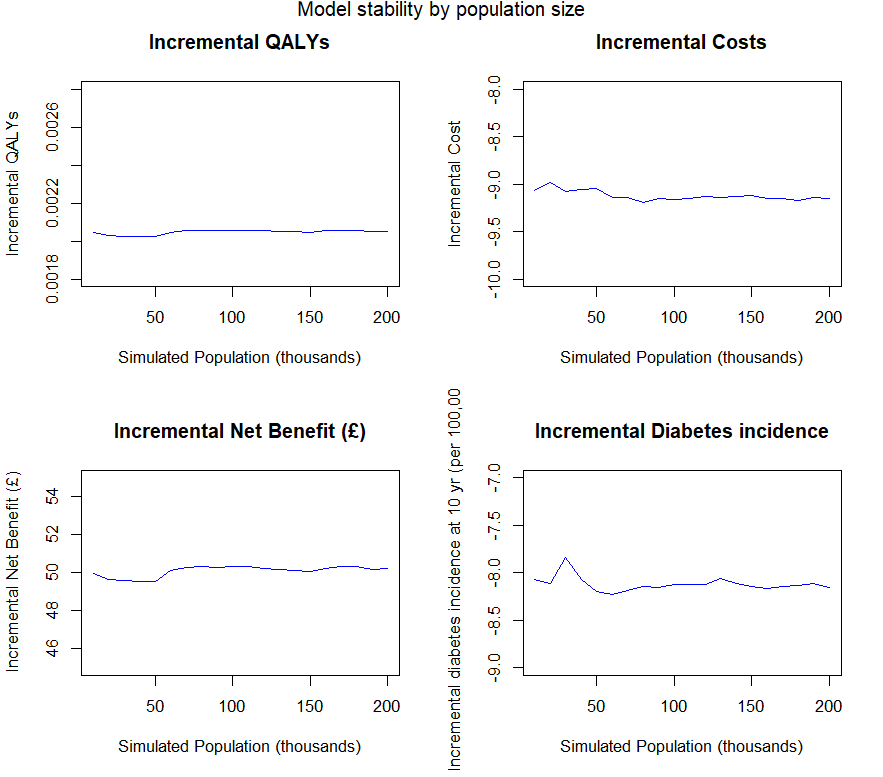

### 11. Model Results

Table S33 reports summary results from sensitivity analyses varying data inputs and key model assumptions.

*Table S33: Results of one-way and structural sensitivity analysis on incremental lifetime cost, QALYs and Net Benefit.*

|  | Description/rationale | Incremental Costs | Incremental QALYs | Incremental Net Benefit |
| --- | --- | --- | --- | --- |
| Base case |  | -£9.16  (-£31.63 to £2.90) | 0.002  (0.001 to 0.005) | £50.23  (-£16.41 to £74.68 |
| SA1: Behavioural effect only | Mean reduction in kcal of -2 per day for 12 months | -£0.19  (-£10.77 to £10.44) | -0.0002  (-0.002 to 0.001) | -£3.88  (-£42.09 to £10.42) |
| SA2: Reformulation effect only | Mean reduction in kcal of -2 per day for lifetime | -£8.93 (-£27.23 to £0.18) | 0.002 (0 to 0.004) | £54.55  (£3.69 to £73.96) |
| SA3: Stratified by sex and occupational socio-economic status | Stratified analysis to capture variation in behavioural effects and out of home purchases by socio-economic status | -£9.71  (-£31.26 to £2.78) | 0.001  (-0.005 to 0.004) | £24.16  (-£104.67 to £63.6) |
| SA4: Alternative menu reformulation effect by IMD | Investigate the impact of source of evidence for reformulation on the results. Evidence from online delivery services suggests a slightly increased effect on calories. | -21.07 (-55.14 to -2.87) | 0.005 (0.002 to 0.007) | £120.84 (£27.86 to £119.76) |
| SA5. Discount rate 0% | Assess the impact of discount rate on overall conclusions. | -£24.97  (-£76.88 to £2.27) | 0.004  (-0.001 to 0.01) | £111.06  (-£22.12 to £142) |
| SA6. Discount rate 5% | Assess the impact of discount rate on overall conclusions. | -£6.44  (-£23.98 to £2.89) | 0.002  (-0.001 to 0.004) | £38.92  (-£18.35 to £62.26) |
| SA7. Alternative evidence from international evidence | Previous modelling studies report a behavioural effect of -47kcal per meal (65), reformulation effect -15kcal per meal (66). | -£17.1 (-45 to -2) | 0.004 (0.002 to 0.007) | £100.00 (27.8 to 105.0) |

| SA8. Alternative evidence from international evidence | Most recent meta-analysis of calorie change report a behavioural effect of -11kcal per meal (68), reformulation effect -15kcal per meal(66). | -£15.3 (-40.2 to -1.1) | 0.004 (0.002 to 0.006) | £93.5 (26.9 to 102.7) |
| --- | --- | --- | --- | --- |
| SA9. Delayed menu reformulation effect | Evidence shows that calories lower in new products. Impact on consumer diets may develop over time as tastes and awareness develops (69). Delay reformulation effects for 3 years. | -£5.77  (-£21.38 to £3.1) | 0.001  (-0.001 to 0.004) | £35.19  (-£18.62 to £58.52) |
| SA10. Add in weight effects on cardiometabolic health | Evidence suggests that weight loss can reduce systolic blood pressure, cholesterol and glycaemia (70). | -£99.31  (-£329.08 to £19.28) | 0.008  (-0.002 to 0.019) | £252.13  (-£67.21 to £187.61) |
| SA11. Alternative calorie to weight model | Convert changes in the energy intakes into changes in bodyweight, based on the Christiansen and Garby energy conservation prediction formula (71) | -£20.09  (-£75.48 to £5.03) | 0.002  (-0.01 to 0.01) | £68.39  (-£230.28 to £156.2) |

### 12. CHEERS Checklist

| **Title** | | | |
| --- | --- | --- | --- |
| Title | 1 | Identify the study as an economic evaluation and specify the interventions being compared. | Y |
| **Abstract** | | | |
| Abstract | 2 | Provide a structured summary that highlights context, key methods, results, and alternative analyses. | Y |
| **Introduction** | | | |
| Background and objectives | 3 | Give the context for the study, the study question, and its practical relevance for decision making in policy or practice. | Introduction |
| **Methods** | | | |
| Health economic analysis plan | 4 | Indicate whether a health economic analysis plan was developed and where available. | Appendix section 1 |
| Study population | 5 | Describe characteristics of the study population (such as age range, demographics, socioeconomic, or clinical characteristics). | Table 1 |
| Setting and location | 6 | Provide relevant contextual information that may influence findings. | Background |
| Comparators | 7 | Describe the interventions or strategies being compared and why chosen. | Methods – Intervention effects |
| Perspective | 8 | State the perspective(s) adopted by the study and why chosen. | Methods – Health Economic Analysis |
| Time horizon | 9 | State the time horizon for the study and why appropriate. | Methods – Health Economic Analysis |
| Discount rate | 10 | Report the discount rate(s) and reason chosen. | Methods – Health Economic Analysis |
| Selection of outcomes | 11 | Describe what outcomes were used as the measure(s) of benefit(s) and harm(s). | Methods – Health Economic Analysis |
| Measurement of outcomes | 12 | Describe how outcomes used to capture benefit(s) and harm(s) were measured. | Methods – Simulation model |
| Valuation of outcomes | 13 | Describe the population and methods used to measure and value outcomes. | Methods – Simulation model |
| Measurement and valuation of resources and costs | 14 | Describe how costs were valued. | Methods – Simulation model |
| Currency, price date, and conversion | 15 | Report the dates of the estimated resource quantities and unit costs, plus the currency and year of conversion. | Appendix section 6 |
| Rationale and description of model | 16 | If modelling is used, describe in detail and why used. Report if the model is publicly available and where it can be accessed. | Methods – Simulation model |
| Analytics and assumptions | 17 | Describe any methods for analysing or statistically transforming data, any extrapolation methods, and approaches for validating any model used. | Appendix |
| Characterising heterogeneity | 18 | Describe any methods used for estimating how the results of the study vary for subgroups. | Methods – Uncertainty Analysis |
| Characterising distributional effects | 19 | Describe how impacts are distributed across different individuals or adjustments made to reflect priority populations. | Methods – Simulation model |
| Characterising uncertainty | 20 | Describe methods to characterise any sources of uncertainty in the analysis. | Appendix Section 3-9 |
| Approach to engagement with patients and others affected by the study | 21 | Describe any approaches to engage patients or service recipients, the general public, communities, or stakeholders (such as clinicians or payers) in the design of the study. | Appendix Section 1 |
| **Results** | | | |
| Study parameters | 22 | Report all analytic inputs (such as values, ranges, references) including uncertainty or distributional assumptions. | Appendix Section 9 |
| Summary of main results | 23 | Report the mean values for the main categories of costs and outcomes of interest and summarise them in the most appropriate overall measure. | Table 3 |
| Effect of uncertainty | 24 | Describe how uncertainty about analytic judgments, inputs, or projections affect findings. Report the effect of choice of discount rate and time horizon, if applicable. | Figure 1 |
| Effect of engagement with patients and others affected by the study | 25 | Report on any difference patient/service recipient, general public, community, or stakeholder involvement made to the approach or findings of the study | Appendix Section 1 |
| **Discussion** | | | |
| Study findings, limitations, generalisability, and current knowledge | 26 | Report key findings, limitations, ethical or equity considerations not captured, and how these could affect patients, policy, or practice. | Discussion |
| **Other relevant information** | | | |
| Source of funding | 27 | Describe how the study was funded and any role of the funder in the identification, design, conduct, and reporting of the analysis | Funding declaration |
| Conflicts of interest | 28 | Report authors conflicts of interest according to journal or International Committee of Medical Journal Editors requirements. | Conflicts of interest statement |

### 13. References

1.	Health Survey for England. NHS Digital2018.

2.	Health Survey for England. NHS Digital2019.

29.	National life tables: England and Wales. In: Statistics OoN, editor. 2023.

30.	Jones KC, Weatherly H, Birch S, Castelli A, Chalkley M, Dargan A, et al. Unit costs of health and social care 2022 manual. 2023.

31.	Excellence NIfHaC. PH38 Preventing type 2 diabetes - risk identification and interventions for individuals at high risk: guidance. National Institute for Health and Care Excellence [Internet]. 2012; NICE public health guidance 38. Available from: <http://guidance.nice.org.uk/PH38/Guidance/pdf/English>.

32.	CG127 Hypertension: costing template. National Institute for Care and Clinical Excellence [Internet]. 2011. Available from: <http://guidance.nice.org.uk/CG127/CostingTemplate/xls/English>.

33.	Poole CD, Tetlow T, McEwan P, Holmes P, Currie CJ. The prescription cost of managing people with type 1 and type 2 diabetes following initiation of treatment with either insulin glargine or insulin detemir in routine general practice in the UK: a retrospective database analysis. Current Medical Research and Opinion. 2007;23(sup1):S41-S8.

34.	British National Formulary. <http://wwwbnforg/> [Internet]. 2024.

35.	guideline NG136 N. Hypertension in adults: diagnosis and management. Methods. 2019.

47.	Jones KC BA. Unit costs of health and social care. PSSRU; 2021 2021.

48.	SSNAP NGC. Sentinel Stroke National Audit Programme: Cost and Cost-effectiveness analysis (Technical Report)2016. Available from: <https://www.strokeaudit.org/SupportFiles/Documents/Health-economics/Health-economic-report-2016.aspx>.

49.	Dolan PG, C; Kind, P.;Williams, A. A social tariff for Euroqol: Results from a general population survey. Discussion Paper 138. 1995;University of York.

50.	Ward S, Lloyd JM, Pandor A, Holmes M, Ara R, Ryan A, et al. A systematic review and economic evaluation of statins for the prevention of coronary events. Health Technol Assess. 2007;11(14):1-iv.

51.	Ara R, Wailoo A. NICE DSU Technical Support Document 12: The use of health state utility values in decision models. 2011 2011.

67.	Mandating calorie labelling of food and drink in out-of-home settings. Impact Assessment (IA). Department of Health and Social Care (DHSC) 2020.

68.	Clarke N, Pechey E, Shemilt I, Pilling M, Roberts NW, Marteau TM, et al. Calorie (energy) labelling for changing selection and consumption of food or alcohol. Cochrane Database of Systematic Reviews. 2025(1).

69.	Grummon AH, Petimar J, Soto MJ, Bleich SN, Simon D, Cleveland LP, et al. Changes in calorie content of menu items at large chain restaurants after implementation of calorie labels. JAMA network open. 2021;4(12):e2141353-e.

70.	Pidd K, Breeze P, Ahern A, Griffin SJ, Brennan A. Effects of weight loss and weight gain on HbA1c, systolic blood pressure and total cholesterol in three subgroups defined by blood glucose: a pooled analysis of two behavioural weight management trials in England. BMJ open. 2025;15(4):e095046.

71.	Christiansen E, Garby L. Prediction of body weight changes caused by changes in energy balance. European journal of clinical investigation. 2002;32(11):826-30.
